# Multi-omics Analysis of Umbilical Cord Hematopoietic Stem Cells from a Multi-ethnic Cohort of Hawaii Reveals the Intergenerational Effect of Maternal Pre-Pregnancy Obesity and Risk Prediction for Cancers

**DOI:** 10.1101/2024.07.27.24310936

**Authors:** Yuheng Du, Paula A. Benny, Yuchen Shao, Ryan J. Schlueter, Alexandra Gurary, Annette Lum-Jones, Cameron B Lassiter, Fadhl M. AlAkwaa, Maarit Tiirikainen, Dena Towner, W. Steven Ward, Lana X Garmire

## Abstract

**Background:** Maternal obesity is a health concern that may predispose newborns to a high risk of medical problems later in life. To understand the intergenerational effect of maternal obesity, we hypothesized that the maternal obesity effect is mediated by epigenetic changes in the CD34+/CD38−/Lin− hematopoietic stem cells (uHSCs) in the offspring. Towards this, we conducted a DNA methylation centric multi-omics study. We measured the DNA methylation and gene expression in the CD34+/CD38−/Lin− uHSCs and metabolomics of the cord blood, all from a multi-ethnic cohort (n=72) from Kapiolani Medical Center for Women and Children in Honolulu, Hawaii (collected between 2016 and 2018).

**Results:** Differential methylation (DM) analysis unveiled a global hypermethylation pattern in the maternal pre-pregnancy obese group (BH adjusted p<0.05), after adjusting for major clinical confounders. KEGG pathway enrichment, WGCNA, and PPI analyses revealed hypermethylated CpG sites were involved in critical biological processes, including cell cycle, protein synthesis, immune signaling, and lipid metabolism. Utilizing Shannon entropy on uHSCs methylation, we discerned notably higher quiescence of uHSCs impacted by maternal obesity. Additionally, the integration of multi-omics data-including methylation, gene expression, and metabolomics-provided further evidence of dysfunctions in adipogenesis, erythropoietin production, cell differentiation, and DNA repair, aligning with the findings at the epigenetic level. Furthermore, we trained a random forest classifier using the CpG sites in the genes of the top pathways associated with maternal obesity, and applied it to predict cancer vs. adjacent normal labels from samples in 14 Cancer Genome Atlas (TCGA) cancer types. Five of 14 cancers showed balanced accuracy of 0.6 or higher: LUSC (0.87), PAAD (0.83), KIRC (0.71), KIRP (0.63) and BRCA (0.60).

**Conclusions:** This study revealed the significant correlation between pre-pregnancy maternal obesity and multi-omics level molecular changes in the uHSCs of offspring, particularly in DNA methylation. Moreover, these maternal obesity epigenetic markers in uHSCs may predispose offspring to higher risks in certain cancers.

## Introduction

Maternal obesity has emerged as a primary health concern during pregnancy, with its prevalence alarmingly increasing. According to a study by the Centers for Disease Control and Prevention, the percentage of women experiencing pre-pregnancy obesity in the United States escalated from 26% to 29% between 2016 and 2019 ^1^. Born to mothers with obesity, higher birth weight is associated with a higher incidence of childhood cancers such as leukemia and neuroblastoma ^2,3^, as well as greater risks of prostate and testicular cancers in men ^4–6^ and breast cancer in women ^7^. Moreover, maternal obesity may have a intergenerational effect and set the stage for increased chronic disease susceptibility later in the adulthood of offspring ^8,9^. The hypothesis of the utero origin of diseases proposes that numerous chronic diseases have their origins in the fetal stage, the earliest phase of human development ^10,11^. Some researchers have speculated higher stem cell burdens in newborn babies born from obese mothers ^12^. Altered hormonal environment and nutrient availability can induce critical changes in fetal stem cells ^13^, which may predispose these cells to malignant transformation, aligning with the idea of the cancer stem cell hypothesis that cancer cells have stem cell-like properties with an uncontrolled self-renewal program ^14–16^. In particular, a study showed increases in cord blood CD34+CD38-stem cell and CD34+ progenitor cell concentrations with maternal obesity ^17^, suggesting that the higher proportions of stem cells in cord blood may make the babies more susceptible to obesity and cancer risks. However, so far little work provides direct molecular links as to how maternal obesity affects cellular function and increases the disease risk in offspring.

To seek answers in this area, we conducted an epigenome-centered multi-omics study to directly pinpoint the effect of maternal obesity in umbilical cord blood hematopoietic stem cells (uHSCs). Epigenetics is chosen as the center of multi-omics integration, as it is both inheritable and susceptible to modification by diseases. Thus, it may serve as a plausible mediator in the transmission of the effects of maternal obesity to offspring. We incorporate gene expression for cord blood stem cells and metabolomics data from the cord blood serum as the downstream readout of epigenetics changes. By elucidating these molecular connections, we provide a systematic understanding of how maternal obesity during pregnancy can influence the multiple types of molecular profiles of newborns. Such knowledge may ultimately help develop early therapeutic interventions at the molecular level to mitigate these intergenerational health risks due to maternal obesity.

## Methods

### Overview of the maternal pre-pregnancy cohort with baby cord blood

In this study, baby cord blood samples from 72 pregnant women (34 obese; 38 non-obese) who delivered at Kapiolani Medical Center for Women and Children in Honolulu, Hawaii (2015-2018) were collected. The study was approved by the Western IRB (WIRB Protocol #20151223). Patients meeting the inclusion criteria were identified from pre-admission medical records with pre-pregnancy BMI ≥ 30.0 (maternal obesity) or 18.5-25.0 (normal weight).

Pregnant women undergoing elected C-sections at ≥37 weeks gestation were included, to minimize confounding events during the labor. Patient exclusion criteria included pregnant women with preterm rupture of membranes, labor, multiple gestations, pregestational diabetes, hypertensive disorders, cigarette smokers, infection of human immunodeficiency virus or hepatitis B virus, and chronic drug use. Demographic and phenotypic information was recorded, including maternal and paternal age, ethnicity, gestational weight gain, gestational age, parity, and gravidity. For newborns, Apgar scores were documented at 1 minute and 5 minutes post-delivery. The Apgar score serves as a comprehensive assessment of a newborn’s health, with a normal range considered to be between 7 to 10 ^18^.

### Sample preparation and methylation profiling

The baby cord blood sample was collected in the operating room under sterile conditions at the time of the C-section (Pall Medical Cord Blood Collection Kit containing 25ml citrate phosphate dextrose). The umbilical cord was first cleansed with chlorhexidine swabs before cord blood collection. The total volume of collected blood was measured and recorded before aliquoting to conical tubes for centrifugation. The tubes were centrifuged at 200g for 10 min, and plasma was collected. The plasma volume was replaced with 2% FBS/PBS. Negative selection reagents were added to the blood and incubated for 20 min (Miltenyi Biotec, Auburn, CA). The cord blood was diluted with an equal volume of 2% FBS/PBS. A 20ml aliquot of the diluted blood was layered over a density gradient (15ml Lymphoprep) and centrifuged at 1200g for 20 min. The top layer containing an enriched population of stem cells was collected, centrifuged at 300g for 8 min, and then washed in 2% FBS/PBS. Red blood cells were lysed using ammonium chloride (9:1) with incubation on ice for 10 min, washed twice, and then resuspended in 100µl of 2% FBS/PBS. Cells were stained with Lineage FITC and CD34 APC for 45 min on ice, washed twice, and then sorted using the BD FacsAria III. Hematopoietic stem cells (CD34+/CD38−/Lin−) were collected and stored at −80°C until DNA/RNA extraction.

DNA and RNA were extracted simultaneously using the AllPrep DNA/RNA extraction kit (Qiagen). DNA purity and concentration were quantified in Nanodrop 2000 and Picogreen assay. Bisulfite conversion of 500 ng DNA was performed using the EZ DNA Methylation kit (Zymo), followed by sample processing for Infinium HumanMethylation450 bead chips (Illumina) according to the manufacturer’s instructions. Bead chips were analyzed at the Genomics Shared Resource at the University of Hawaii Cancer Center.

### Bulk RNA sequencing

A total of 50 RNA samples were prepared for bulk RNA Sequencing. RNA concentration and RIN score were assayed using Nanodrop 2000 and Agilent Bioanalyzer. A total of 200 ng of high-quality RNA (RIN≥7) was subjected to library construction (polyA) and sequenced on HS4000 (2×100) at the Yale Center for Genome Analysis, Connecticut to obtain at least 25M paired reads per sample.

### Methylation data pre-processing

The overall preprocessing workflow is shown in **Supplemental Figure 1**. R version 3.6.3 was used for all analyses. As the first step of quality assessment, sex chromosome methylation patterns were analyzed to check for potential sex mismatches between reported and inferred sex using the getSex() function in minfi^19^. No samples with discrepancies between reported and inferred sex were identified or flagged for exclusion **Supplemental Figure 2A**. Raw intensity data (.idat) were extracted using the ‘ChAMP’ package (version 2.16.2) in R with *champ.load()* function ^19–22^. For the filtration step, background controls were subtracted from the data, and raw data that did not pass detection P-value of 0.05 were removed. CpG sites whose probes had known underlying SNPs and association with XY chromosomes were removed from analysis due to potential confounding. The quality controls included checking the raw density distribution, multi-dimensional scaling (MDS) plot, and median intensity values of methylated and unmethylated probes to identify potential outliers or poorly performing samples (**Supplemental Figure 2B-D**). After BMIQ normalization using *champ.norm()* function ^23^, the batch effect (including slide and array) due to non-biological technical variation caused by experiment handling was removed using the ComBat function in the ChAMP package, confirmed by the singular value decomposition (SVD) plot (**Supplemental Figure 2E**). A total of 1,992 cross-hybridizing probes were removed using the probe list from ExperimentHub (query id ‘EH3129’) as reported in Chen et al. 2013 ^24^. For each CpG site, the methylation score was initially calculated as the beta value, a fluorescence intensity ratio between 0 and 1. To reduce the heteroskedasticity for downstream statistical analysis, the M-values were transformed from beta-values using lumi (ver 3.1.4) in R ^25–28^. A total of 408,773 CpG sites remained for downstream analysis after probe filtering, quality control, normalization, batch correction, and cross-hybridizing probes removal.

### Source of variation analysis and confounding adjustment

To eliminate potential confounding factors of pre-pregnant maternal obesity among the 13 clinical factors, we conducted a source of variation analysis with a collection of ANOVA tests to identify the clinical factors that significantly contribute to the methylation level variation, as done before ^29,30^. The variables with F statistics greater than 1 (the error value) were determined as confounders and subjected to confounding adjustment. These factors include the baby’s sex, net weight gain during the pregnancy, maternal age, maternal ethnicity, paternal ethnicity, gravidity, and gestational age. To adjust for confounding effects, a multivariate regression model is built using the ‘limma’ package to fit methylation M values of each CpG site, using the confounding factors above. The remaining residuals on the M values were considered to be confounding adjusted for the subsequent bioinformatics analysis of DNA methylation. To assess the bias and inflation in the differential methylation findings, we used Bayesian method “bacon” to calculate the genomic inflation (lambda) values before and after the confounder adjustment ^31^. Additional surrogate variable analysis (sva) and randomly shuffled null lambda calculation were performed to determine the need for inflation adjustment ^32^. The null model lambda was 0.96. No surrogate variables were identified for correction in the adjusted model. Thus the observed inflation reflects the true biological signal rather than systematic bias, and no further inflation correction was performed on this confounder adjusted model.

### Bioinformatics analysis of differential methylation (DM)

A moderated t-test from the ‘limma’ R package (version 3.42.2) ^33^ was used for detecting DM CpG sites between healthy controls and cases with M values. The p-values were adjusted for multiple hypotheses testing using Benjamini-Hochberg FDR. CpG sites with FDR <0.05 were considered statistically significant. To minimize the effect of the gestational age, CpG sites located within the gestational-age-related differentially methylated regions (DMR) were removed. A total of 130 DMRs related to gestational age were identified using linear regression analysis performed with bumphunter ^34^ across eight public datasets including a total of 248 patients.: GSE31781 ^35^, GSE36829 ^36^, GSE59274 ^35,37^, GSE44667 ^38^, GSE74738 ^39^, GSE49343 ^40^, GSE69502 ^41^, and GSE98224 ^42,43^. The complete list of DMRs was included in **Supplemental Table 1**. Hypermethylation and hypomethylation states were defined by the values of log2 Fold Change (log2FC) of M values in cases compared to controls: hypermethylation if bigger than 0, and hypomethylation if less than 0. Corresponding genes and feature locations of these differential CpG sites were annotated using IlluminaHumanMethylation450kanno.ilmn12.hg19 (ver 0.6.0) ^44^.

### KEGG pathway enrichment analysis

‘gometh’ function from R package “missMethyl” (version 1.26.1) ^45–48^ was used for KEGG pathway enrichment ^49–51^ with DNA methylation data. DM sites were used for pathway enrichment within five supergroups from KEGG pathways: Metabolism, Genetic Information Processing, Environmental Information Processing, Cellular Processes, and Organismal Systems. Pathways with adjusted p-values less than 0.05 were considered significant. Pathway scores for protein pathways (KEGG: Transcription, Translation, Folding, sorting and degradation) and immune pathways (KEGG: Immune system) were calculated with averaged beta values from the promoter region CpG sites. To validate the enrichment of significant CpGs in specific pathways, we used the hypergeometric test, which calculates the probability of observing k or more significant CpGs in a pathway by chance, given the total CpGs on the Illumina array (N), the total CpGs in the pathway (K), and the total significant CpGs identified in our study (n). The formula is: 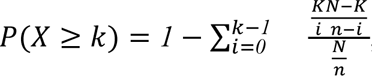, where 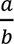 represents the binomial coefficient.

### Weighted co-expression network analysis

Firstly, we adjusted all beta values with clinical confounders, then summarized the DM CpG sites at the gene level by averaging the beta values in the promoter regions (those in the TSS200 and TSS1500 promoter regions). Next, we transformed adjusted beta values to adjusted M values for the downstream adjacency matrix construction. We used adjusted M values for the weighted gene co-expression network analysis (WGCNA) with R package ‘WGCNA’ (version 1.70-3) ^52,53^. The soft threshold for the weighted adjacency matrix with an adjusted R^2^>0.8 was 7. The topological overlap matrix was constructed for hierarchical clustering. Modules were identified by the dynamic tree-cut algorithm. The networks were exported to Cytoscape with an edge weight greater than 0.03 in each module. The genes with the highest betweenness and degree in the WGCNA network were identified as the hub genes for different modules.

### Protein-protein interaction network analysis

For the protein-protein interaction (PPI) network analysis, DM genes are used as the inputs and were mapped on the STRING database (version 10) ^54^. Significantly functionally associated protein pairs were identified using PANDA (Preferential Attachment based common Neighbor Distribution derived Associations) (version 0.9.9) ^55^. KEGG pathways associated with these protein pairs (in terms of genes) were found using PANDA. The bipartite network graph was visualized using Cytoscape (version 3.8.1) ^56^.

### Stemness score computation

The stemness score was based on Shannon entropy and scaled plasticity, as proposed previously ^57^. Shannon entropy has been widely applied in developmental biology, particularly in stem cell research ^58–60^. The formulas are shown below:

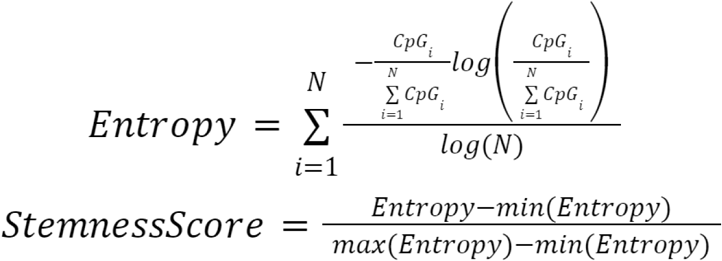

N is the total number of CpG sites. CpG is represented by the beta value on each CpG probe. The stemness score was calculated for all samples using all remaining 408,773 CpG sites after the preprocessing. A Wilcoxon rank test was performed between the stemness scores of the healthy and maternally obese groups.

### Bulk RNA-seq data processing

The Illumina universal adapter regions of raw RNA-seq data were first trimmed using BBMap (version 38.91) ^61^. All raw sequences passed the quality control using fastqc (version 0.11.8) ^62^. The trimmed .fastq files were aligned by STAR (version 2.7.0f) ^63^ to the human Ensembl genome (Homo_sapiens.GRCh38.dna.primary_assembly.fa) and Ensembl annotation (Homo_sapiens.GRCh38.94.gtf). The gene expression counts were calculated using featureCount ^64^ from Subread (ver 1.6.4) ^65^.

### Differential expression (DE) of RNA-Seq data

The limma voom transformation was used on RNA-seq data to model the mean-variance relationship of the log2 counts ^66^, supporting the empirical Bayes analysis pipeline in limma (**Supplemental Figure 3A**). Source of variance analysis was performed to find the clinical confounders with ANOVA tests. The significant confounders included: Maternal_Age, baby sex, hemoglobin, sample group, net weight gain, maternal ethnicity, gravidity and parity (**Supplemental Figure 3B**). The statistically significant DE genes between healthy controls and maternally obese cases were found with confounder adjustment using the ‘DESeq2’ (version 1.26.0) ^67^ and ‘limma-voom’ function from ‘limma’ package ^33^. The p-values were adjusted for multiple hypotheses testing using BH adjustment. No significant differential genes were found with adjusted p-values less than 0.05.

### Correlation analysis between bulk RNA-seq and methylation data

A subset of 47 patients have done both methylation and RNA-seq assays. Pearson correlation coefficients (PCC) were calculated between gene expression and methylation beta values from the promoter regions, among the same patients. As mostly a negative correlation between gene expression and DNA methylation in the promoter region is expected, genes with a high negative correlation (PCC<-0.2) were used for pathway enrichment using TOPPFUN ^68–70^. Top genes of interest were selected with the absolute value Fold Change>1.5 in gene expression and gene-methyl correlation <-0.3 for hyper- and hypo-methylated CpG sites.

### Metabolomics analysis

Metabolomics data were acquired from a previously published study involving 87 patients in the same cohort from three batches (metabolomics workbench study ID ST001114) ^71^. Targeted metabolites were generated with LC-MS, and untargeted metabolites were generated with GC-MS. After the removal of compounds missing in more than 10% of samples, a total of 185 metabolites remained, including 10 amino acids (AA), 40 acylcarnitines (C), 35 acyl/acyl phosphatidylcholines (PC aa), 38 acyl/alkyl phosphatidylcholines (PC ae) and 62 untargeted metabolites. The raw metabolite data were log-transformed, standardized, normalized using variance stabilization normalization (VSN), and batch corrected with ComBat function in sva pacakge^72^. Differential metabolites were identified by limma, with clinical confounders adjustment.

### Multi-omics integration on metabolomics, epigenomics, and transcriptomics

A subset of 42 patients have the matched methylation, gene expression, and metabolomics data. We applied multi-omics integration with Data Integration Analysis for Biomarker discovery using Latent cOmponents (DIABLO) implemented in the mixOmics package ^73^. DIABLO finds the correlated consensus latent variables among different omics in the supervised manner. Top DIABLO features for each omic were selected based on the loading values. We integrated the pathway-level methylation, gene, and metabolite interaction using pathview ^74^.

### Evaluation of maternal pre-pregnancy obesity biomarkers in cancer prediction

We collected Infinium HumanMethylation450 data for a total of 14 cancer datasets (adjacent normal samples > 10): BLCA, BRCA, COAD, ESCA, HNSC, KIRC, KIRP, LIHC, LUAD, LUSC, PAAD, PRAD, THCA, UCEC from The Cancer Genome Atlas (TCGA data portal: https://portal.gdc.cancer.gov/). In total, 6428 samples were obtained, consisting of 5715 tumor samples and 713 adjacent normal tissues.

To build the obesity classification model with maternal obesity biomarkers, we selected 61 hypermethylated CpG sites from the promoter regions of the genes involved in the top five significant pathways based on the missMethyl KEGG enrichment results. This includes cell cycle, ribosome, nucleocytoplasmic transport, ribosome biogenesis in eukaryotes, and mTOR signaling pathway. We split the 72 maternal obesity and control samples at a ratio of 80/20 with 5-fold cross-validation, then constructed a series of classification methods using the *lilikoi* R package, where Random Forest (RF) was the winning model ^75,76^. Next, we applied this RF obesity model on 14 TCGA dataset to perform cancer/normal sample prediction. We report accuracy, balanced accuracy, and F1 score for model performance as done before ^75^.

## Results

### Overview of study design and cohort characteristics

This study aims to investigate the intergenerational effect of pre-pregnancy maternal obesity on offspring. A total of 72 patients who elected to deliver full-term babies through C-sections were recruited from Kapiolani Medical Center for Women and Children in Honolulu, Hawaii from 2016 to 2018. This cohort reflects the multi-ethnic population character of Hawaii, including Asian (N=29), Caucasian (N=15), and Native Hawaiian and Pacific Islanders or NHPIs (N=28). Among them, 38 deliveries are in the healthy control group and 34 are cases with pre-pregnancy maternal obesity. We excluded natural virginal births, to avoid its potential confounding effect on multi-omics profiles. We also carried out stringent recruitment selection criteria, including matching the mothers’ ages as much as possible, as well as similar net gestational weight gain to minimize its confounding effect over maternal pre-pregnancy maternal obesity. The overall study design is shown in **Figure 1**. Briefly, upon collecting the blood samples, umbilical cord blood hematopoietic stem cells (uHSCs) were enriched by FACS sorting with CD34+CD35−LIN− markers (see **Methods**). We extracted DNA and RNA from these uHSCs for Illumina 450k array based DNA methylation and bulk RNA-Seq sequencing respectively. The plasma from these cord blood samples was subject to untargeted metabolomics assays using GC-MS and targeted metabolomics assays using LC–MS ^71^. Given the rationale that DNA methylation could be the mediator for exerting the intergenerational effect of maternal obesity ^77,78^, we carried out multi-omics data integration analysis in the DNA methylation-centric manner.

**Figure 1.**
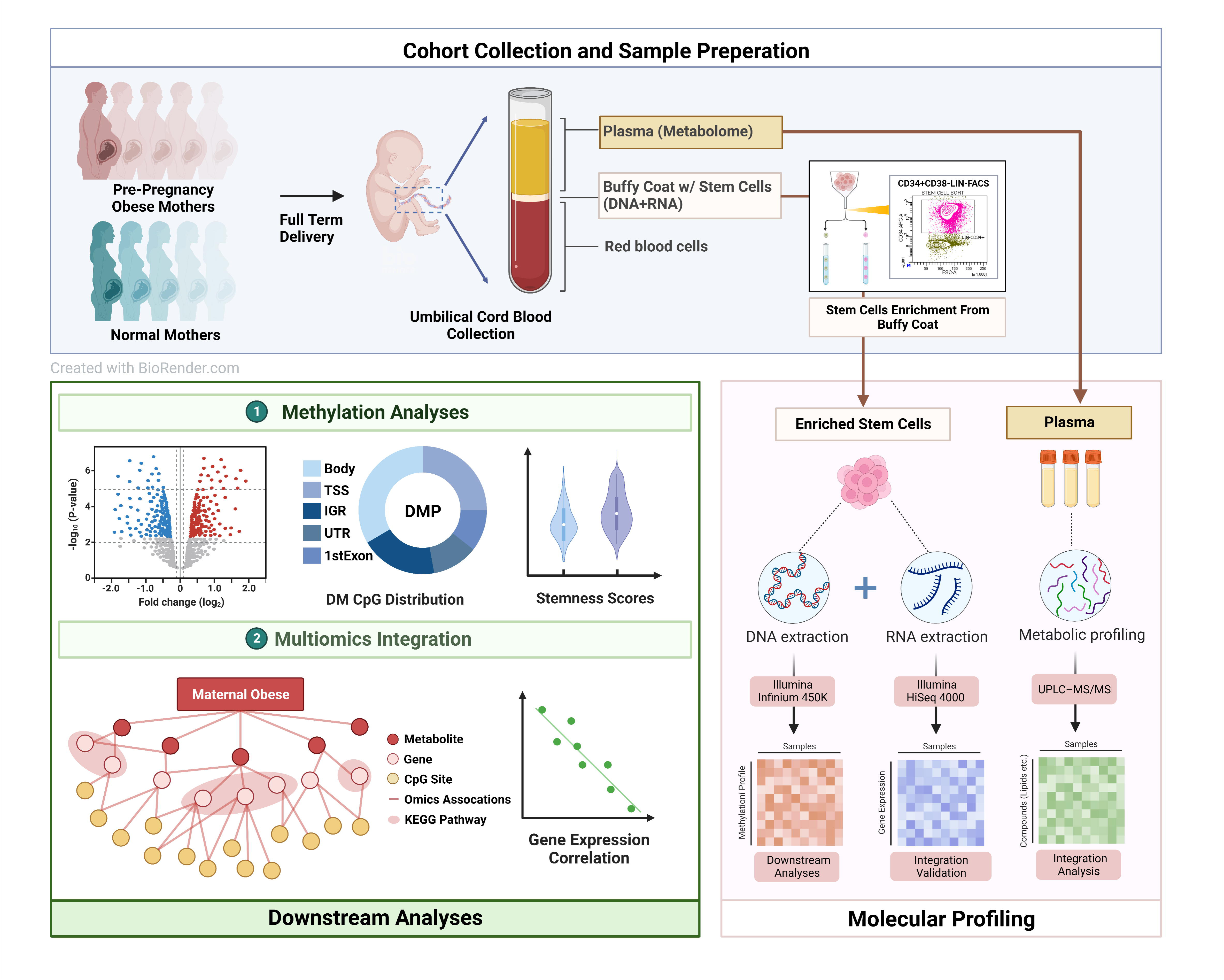
Overview of the study design and analysis. In the preparation step, cord blood plasma samples are collected for metabolome profiling and stem cell sorting. DNA and RNA extraction assays are performed on the enriched stem cells for the methylation and RNA-seq analyses. Downstream analyses are mainly focused on the methylation data. Bulk RNA-seq data were used for validations for methylation discoveries. (Created with BioRender.com)

The demographic details and clinical information of these patients are summarized in **Table 1**. The distributions of the most representative variables are shown in **Figure 2**. Among categorical demographic variables, the distribution of baby sex had no statistical difference between obese and health groups, whereas the ethnicity distributions among mothers and fathers, parity and gravidity are statistically different (P<0.05) between the two groups (**Figure 2A-2E**). Besides maternal pre-pregnancy BMI, other maternal parameters such as maternal age, gestational week, net weight gain and hemoglobin are also not statistically significantly different between the two groups per study design (**Figure 2F-2I**, **Table 1**). While mothers of Asian ethnicity are the majority in the control group, NHPIs account for the majority of the maternal-obese group, revealing the health disparity issue known in the state of Hawaii ^79^. Moreover, the control group has lower parities and gravidities, compared to the cases. Babies born to obese mothers show significantly higher (P<0.05) body weights compared to the control group, as expected ^80^. Other parameters including the baby gender, head circumference, body length, and APGAR score at 5 min after birth are not statistically significantly different between case and control groups (**Figure 2J-2M**).

**Figure 2.**
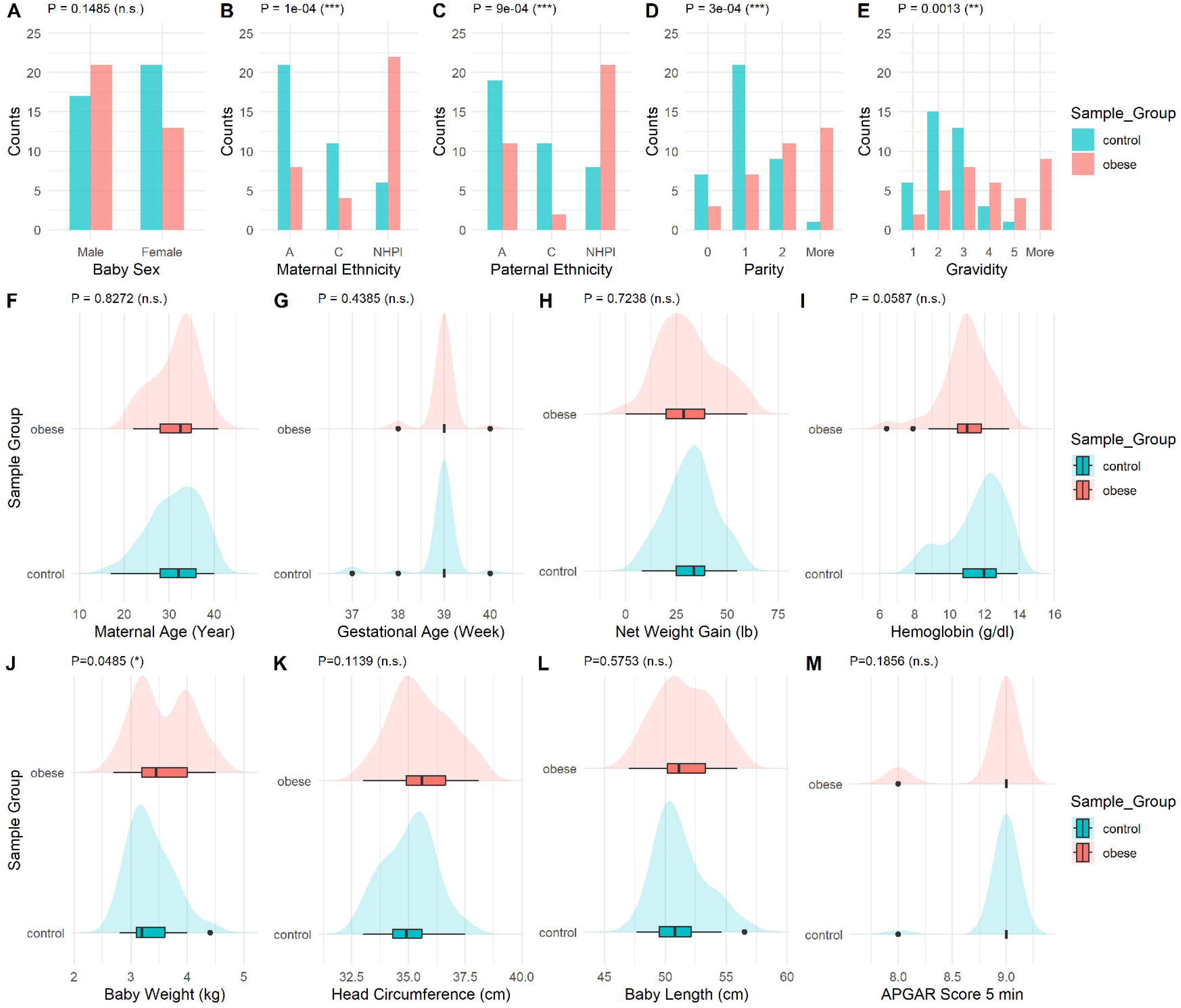
Mother and newborns statistics of the multi-ethnic cohort from Hawaii. (**A-E**) Categorical variables including baby sex, maternal ethnicity, paternal ethnicity, parity and gravidity between control and obese groups are shown in the barplots. P-values using Chi-square test are annotated comparing control and obese groups. **(F-I**) The distributions of maternal age, gestation age, maternal net weight gain during pregnancy, and maternal hemoglobin between control and obese groups are compared. Mean and standard deviation are shown in boxplot. P-values using t-test are annotated. (**J-M**) The distributions of baby weight, baby head circumference, baby length, and APGAR score after 5 minutes of delivery between control and obese groups are compared. Mean and standard deviation are shown in boxplot. P-values using t-test are annotated.

**Table 1.** Summary statistics of the study cohort.

|  |  | Control (n=38) | Case (n=34) |
| --- | --- | --- | --- |
| Maternal Age |  | 31.3±5.6 | 31.6±4.9 |
| Gestational Week |  | 38.9±0.5 | 39.0±0.3 |
| Net Weight Gain |  | 32.0±11.6 | 30.9±14.6 |
| Hemoglobin |  | 11.6±1.6 | 11.0±1.4 |
| Maternal Ethnicity | Asian | 21 | 8 |
|  | Caucasian | 11 | 4 |
|  | NHPI | 6 | 22 |
| Paternal Ethnicity | Asian | 19 | 11 |
|  | Caucasian | 11 | 2 |
|  | NHPI | 8 | 21 |
| Baby Sex | Female | 17 | 21 |
|  | Male | 21 | 13 |
| Parity | 0 | 7 | 3 |
|  | 1 | 21 | 7 |
|  | 2 | 9 | 11 |
|  | More | 1 | 13 |
| Gravidity | 1 | 6 | 2 |
|  | 2 | 15 | 5 |
|  | 3 | 13 | 8 |
|  | 4 | 3 | 6 |
|  | 5 | 1 | 4 |
|  | More | 0 | 9 |

Demographic and clinical statistics are reported for the control and maternally obese groups.

### Global hypermethylation pattern revealed by CpG level methylation analysis

Quality control of methylation data showed no significant sample outliers and no remaining batch effect after ComBAT correction (**Supplemental Figure 2D**). For scientific rigor, it is critical to adjust for confounding in DNA methylation association analysis ^81^. Thus we performed the source of variance (SOV) analysis on the beta values of the DNA methylation with respect to physiological and phenotypic information, in order to assess potential confounding factors systematically ^29,30,81^. As shown in **Figure 3A**, marginal F-statistics in the SOV analysis show that the dominating contribution to DNA methylation variation is maternal pre-pregnancy obesity status, confirming the quality of the study design which aimed to minimize other confounders’ effect. The other minor confounding factors include baby sex, maternal age, maternal ethnicity, net weight gain during pregnancy, paternal ethnicity, gravidity, and gestational age (F-statistics>1). After adjusting these factors by linear regression, all have reduced F-statistics of less than 0.5 (**Figure 3B**) except maternal pre-pregnancy obesity, confirming the success of confounding removal. The quantile-quantile (QQ) plot and genomic inflation factor were used to assess the confounder adjusting model (**Supplemental Figure 4**). A decrease in genomic inflation factor (lambda) was observed with adjustment of confounders. Although the adjusted model had lambda of 1.28, no surrogate variables were identified for correction in the adjusted model. Therefore the observed inflation reflects mostly the true biological signal, and no further inflation correction was performed.

**Figure 3.**
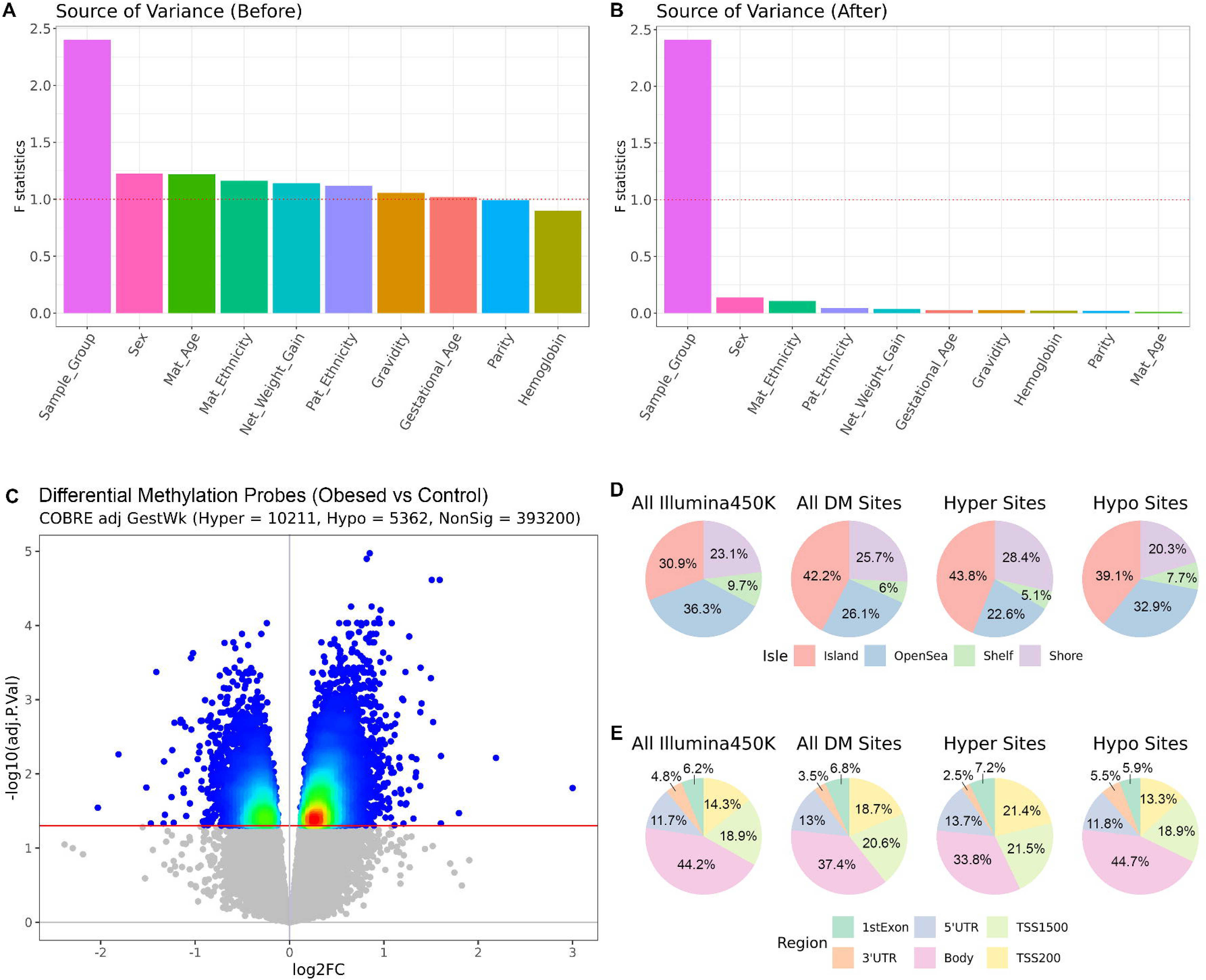
DNA methylation analysis on uHSCs. (**A-B**) Source of variance plot before and after confounding adjustment. F-statistics are reported for each clinical factor. F statistics greater than 1 are considered to have confounding effects in addition to the case/control difference due to pre-pregnancy maternal obesity. (**C**) Volcano plot of −log(BH adjusted p-values) against logFC. The cutoff line for adjusted p-value < 0.05 is shown as the red horizontal line. The hyper/hypo threshold is shown as a blue vertical line where logFC=0. Non-significant methylation CpG sites after the differential analysis were shown in gray. Significant CpG sites are colored. (**D-E**) Normalized location distribution of differentially methylated CpG sites according to their CpG features in terms of isle regions and gene regions based on the chip annotation.

Next, we conducted differential methylation (DE) analysis on the confounding adjusted DNA methylation data (**Methods**). We observed a global hypermethylation pattern in pre-pregnancy obese samples, with 10,211 hypermethylated vs. 5,362 hypomethylated CpG sites (**Figure 3C**). The top 20 differentially hypermethylated and hypomethylated CpG sites are reported in **Table 2**, respectively. These CpG sites are related to a wide variety of biological functions, including inflammation (CD69, ADAM12), transcription factors (ZNF222, HMGN4, LHX6, TAF3), proliferation and apoptosis (HDAC4, DHRS4, LRCH3, SAFB2, CRADD, EBF3, PRKAR1B). Some top DM CpG sites are directly associated with obesity, including HDAC4 ^82^ and PLEC1^83^.

**Table 2.**
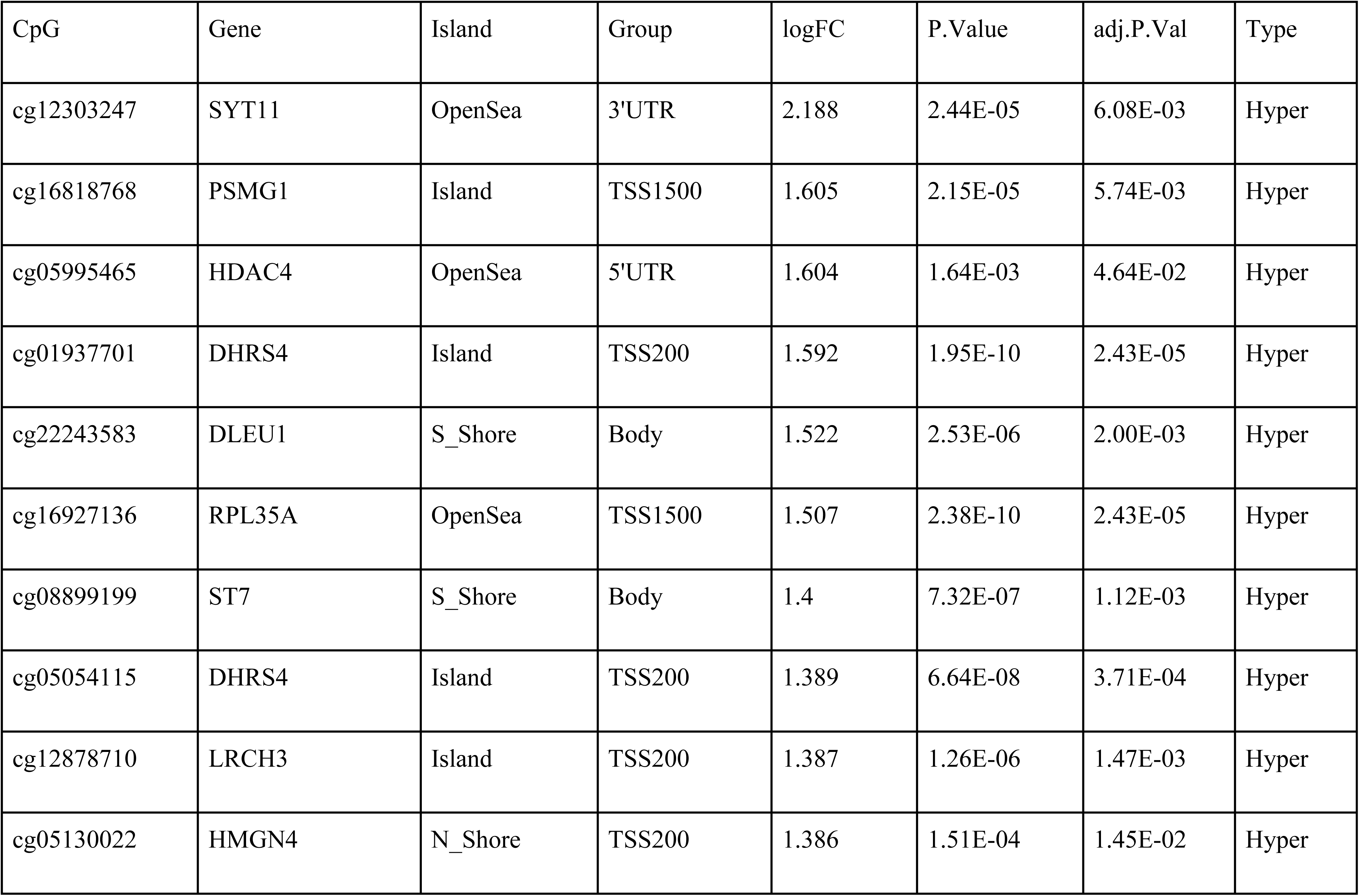
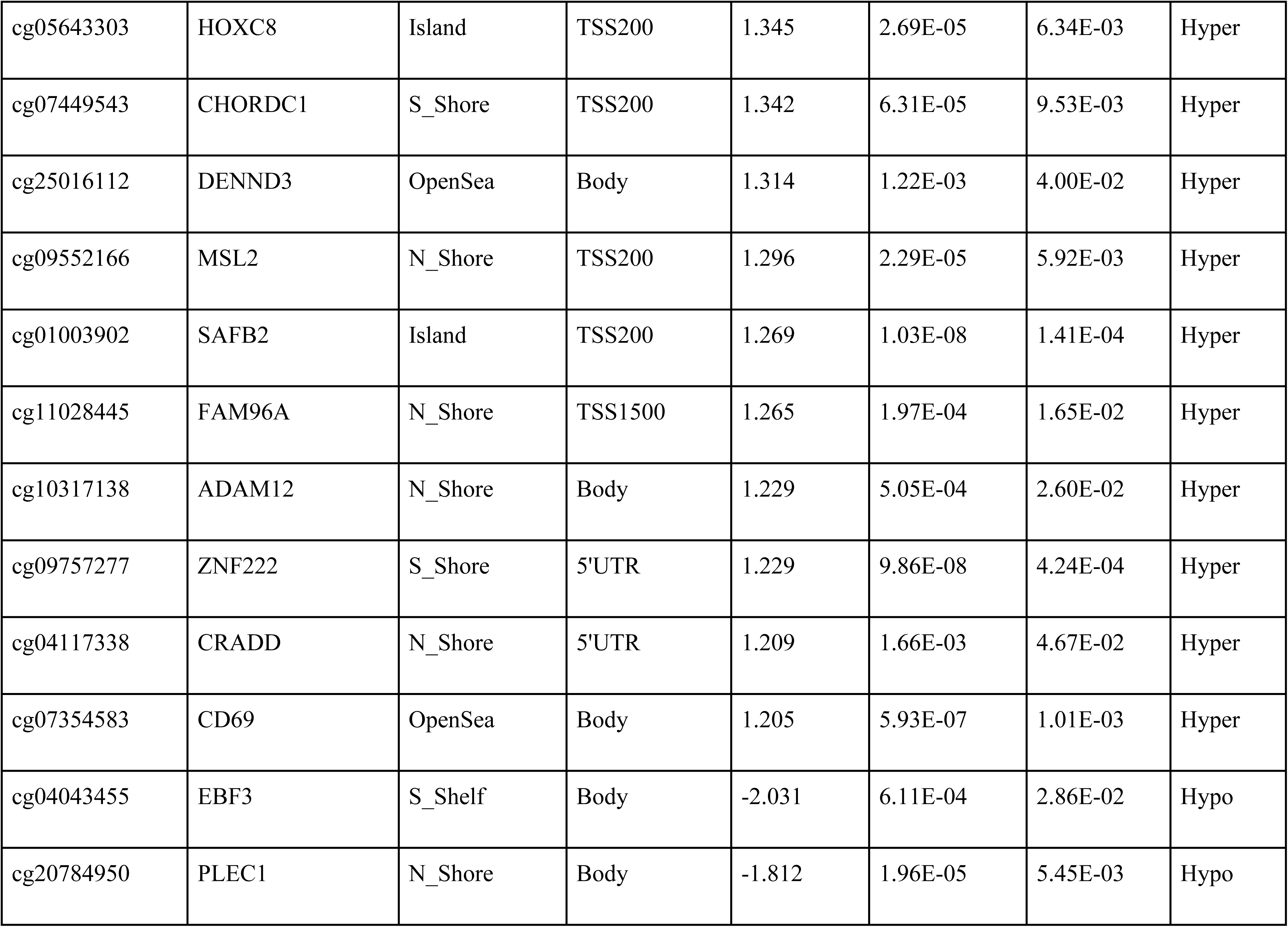
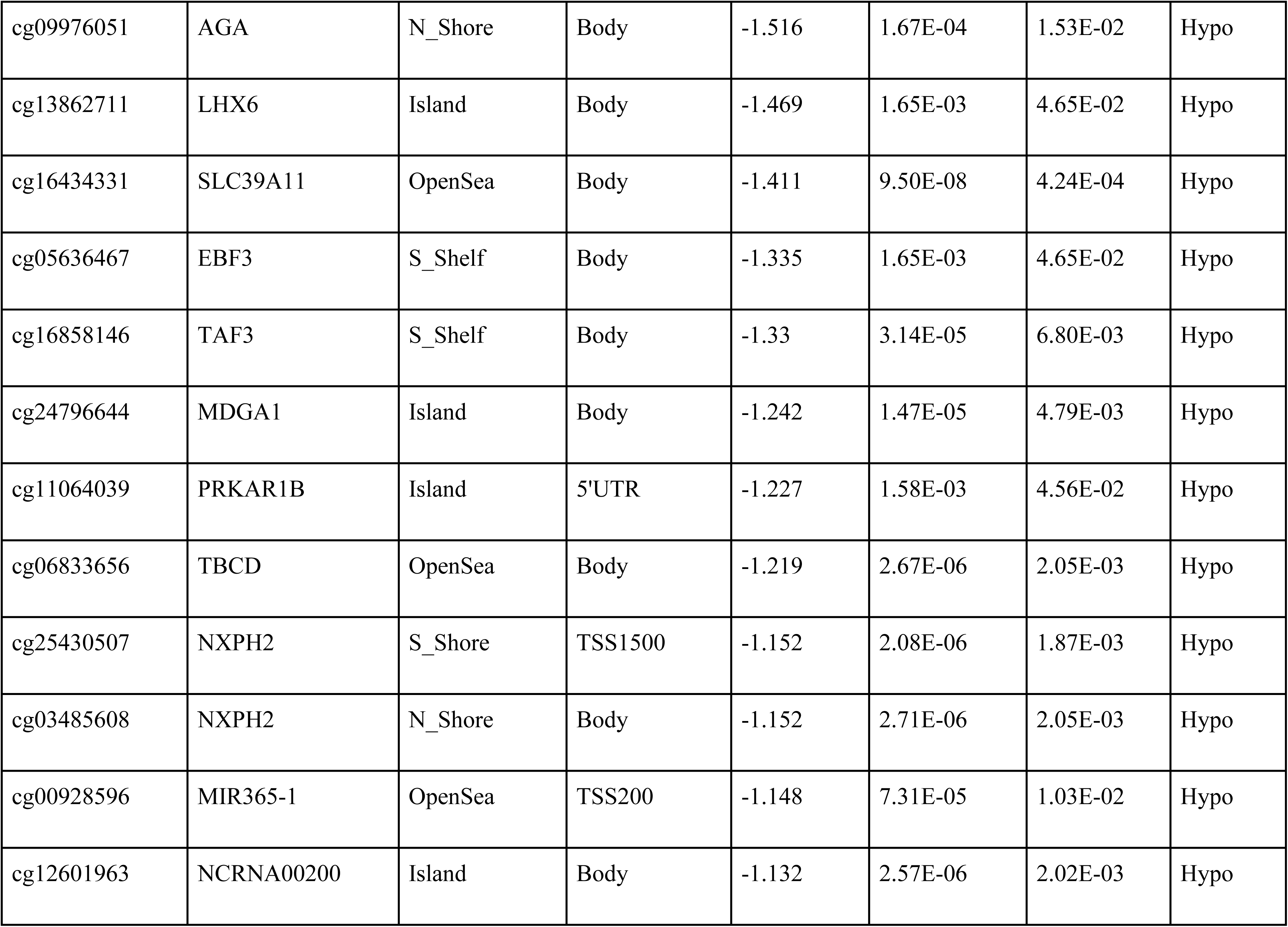
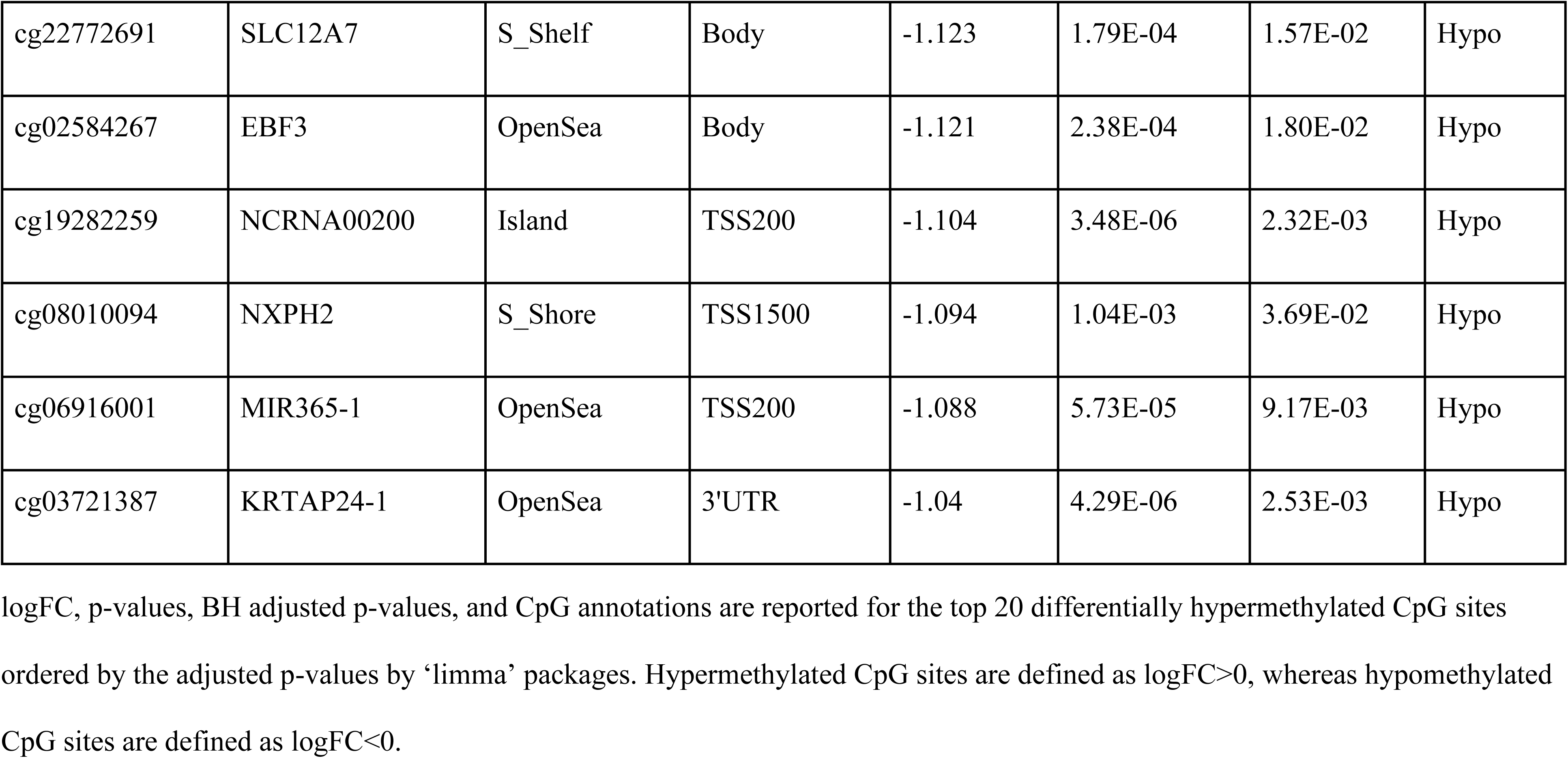
Top 20 hypermethylated CpG sites and top 20 hypomethylated CpG sites.

We further examined the distributions of these differentially methylated sites, relative to the CpG island regions and promoter proximity (**Figure 3D-E**). A big fraction (42.2%) of the DM sites are located in CpG islands ^84,85^, significantly higher than that from the Illumina 450K annotation (P<2E-16). CpG islands are more frequent in the hypermethylated sites (43.8%) than in the hypomethylated sites (39.1%), which is consistent with the global hypermethylation pattern. Relative to gene localization, DM sites are most frequent (39.3%) in the promoter regions (including 18.7% and 20.6% in TSS200 and TSS1500 respectively) as expected.

### Functional analyses reveal the association between maternal obesity and cell cycle, immune function and metabolic changes in the cord blood of offspring

To investigate the biological functions related to the epigenome alternation, we conducted systematic analysis of DM sites employing multiple methods: KEGG pathway enrichment analysis, Weighted Gene Co-expression Network Analysis (WGCNA), and Protein-Protein Interaction (PPI) network analysis.

KEGG pathway enrichment analysis on hypermethylated CpG sites identified five significant pathways with hypergeometric FDR<0.05 (**Figure 4A**), including the cell cycle, ribosome, nucleocytoplasmic transport, ribosome biogenesis in eukaryotes, and mTOR signaling pathway. Cell cycle, ribosome, and nucleocytoplasmic transport pathways are essential to normal cell functioning. mTOR signaling pathway coordinates the nutrient-mediated metabolism, immune responses and cell cycle progression, and dysregulation of mTOR could lead to various diseases such as cancer and obesity ^86^. There was no significantly enriched pathway emerging from hypomethylated CpG sites. The maternally obese group shows significantly higher methylation levels in KEGG protein synthesis and immune system pathway collections compared to the control group, indicating repression in immune response as well as translation and protein synthesis (**Figure 4B-C**). Similarly, we further explored the differential potential, or stemness, of umbilical cord Hematopoietic Stem Cells (uHSCs). We first confirmed the homogeneity of uHSCs by single-cell RNA sequencing UMAP plot (**Supplemental Figure 5**). We calculated the cell stemness scores using the DNA methylation beta values similar to others ^87^. uHSCs derived from the maternally obese group exhibit significantly elevated stemness scores (P<0.01) in comparison to the control group (**Figure 4D**), confirming the results in KEGG pathway analysis.

**Figure 4.**
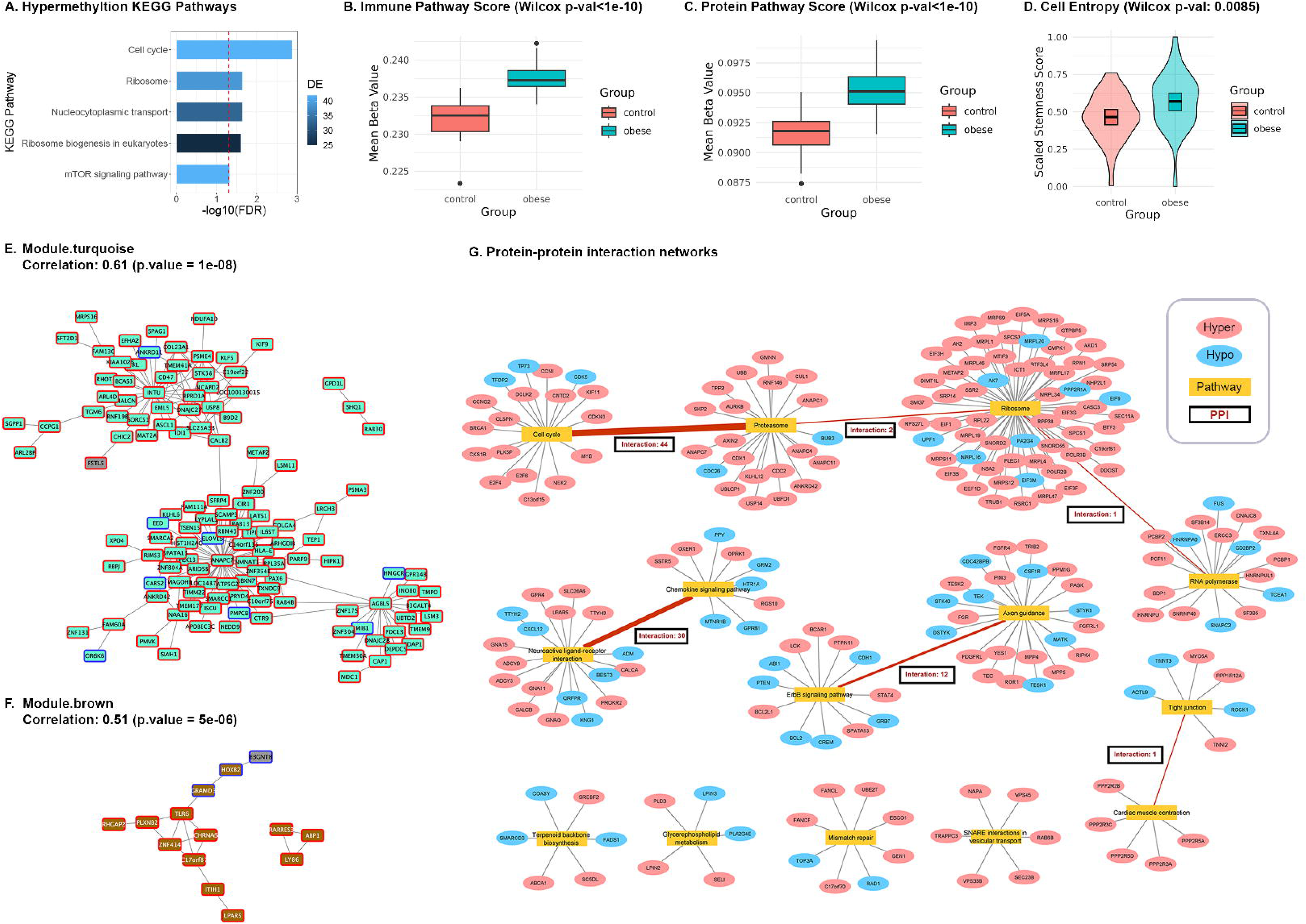
Pathway and network analysis. (**A**) KEGG pathway enrichment for hypermethylated CpG sites from promotor region. Enriched KEGG pathway names, adjusted p-values (-log10 transformed), and the size of enriched gene list are reported for CpG sites from TSS200+TSS1500 regions. The red dotted line shows the threshold cutoff for FDR at −log10(0.05). (**B-C**) Violin plots of averaged beta values for KEGG protein pathway collection and immune pathway collection with Wilcoxon P-values. (**D**) Violin plots of cell entropy scores between control and obese groups with Wilcoxon P-values. (**E-F**) WGCNA network analysis results. WGCNA modules are shown for both the control and the obese group. The top two modules with largest degrees are turquoise and brown modules. Each node represents a gene. Genes co-expressed in each module are annotated. (**G**) Protein-protein interaction (PPI) network. Bipartite graphs represent enriched KEGG pathways and associated genes with significant PPIs. Red nodes represent genes with hypermethylated CpG sites. Blue nodes represent genes with hypomethylated CpG sites. Yellow nodes represented the enriched KEGG pathways. Number of inter-pathway PPIs are annotated in the rectangular boxes.

Next, we applied WGCNA to cluster co-regulation of gene-level methylation, by averaging CpG sites to affiliated genes (see **Methods**). Five co-expression modules are identified, using the M-values adjusted for clinical confounders (**Supplemental Figure 6A)**, and all modules show positive correlations with maternal obesity except one. The largest turquoise module (**Figure 4E**) is related to cell cycle, protein synthesis, and transport and vesicle trafficking pathways through pathway enrichment analysis. Some hub genes in this module are identified, including INTU, ANAPC7, and AGBL5. These genes were reported essential for maintaining cell polarity (INTU)^88^, proliferation (ANAPC7) ^89^ and glycemic control (AGBL5) ^90^. The brown module (**Figure 4F**) is enriched with immune response pathways, in which TLR6 is identified as a hub gene. The other yellow module is related to ion homeostasis, and the gray module is related to the p53 pathway, apoptosis, cell senescence, and ER stress (**Supplemental Figure 6B)**. The only negatively correlated blue module is associated with axon guidance and VEGF signaling pathway (**Supplemental Figure 6B**).

Furthermore, we examined the PPI network, using the gene-level DNA methylation as surrogates (**Figure 4G**). The PPI analysis identifies 14 unique pathways (FDR < 0.05) predominantly associated with hypermethylated CpG sites in the TSS200 and TSS1500 regions. The top five largest pathways included ribosome, proteasome, cell cycle, axon guidance, RNA polymerase, and neuroactive ligand-receptor interaction. Taken all three types of systematic analyses together, cell cycle, immune function and protein synthesis are ubiquitously highlighted, suggesting that these biological functions in cord blood stem cells are negatively impacted by maternal obesity.

### Multi-omics analysis reveals disruptions in cell cycle and metabolic pathways

To systematically investigate the epigenetic, transcriptomic, and metabolomic alterations induced by maternal obesity, we performed multi-omics integration analysis on this cohort. We employed DIABLO, a supervised integration method that extracts features associated with maternal obesity, based on the correlations in the embedding space ^73^. **Figure 5A-C** shows that methylation data provide the clearest separation between obese and control groups, confirming the value of the earlier DNA methylation-centered analysis.

**Figure 5.**
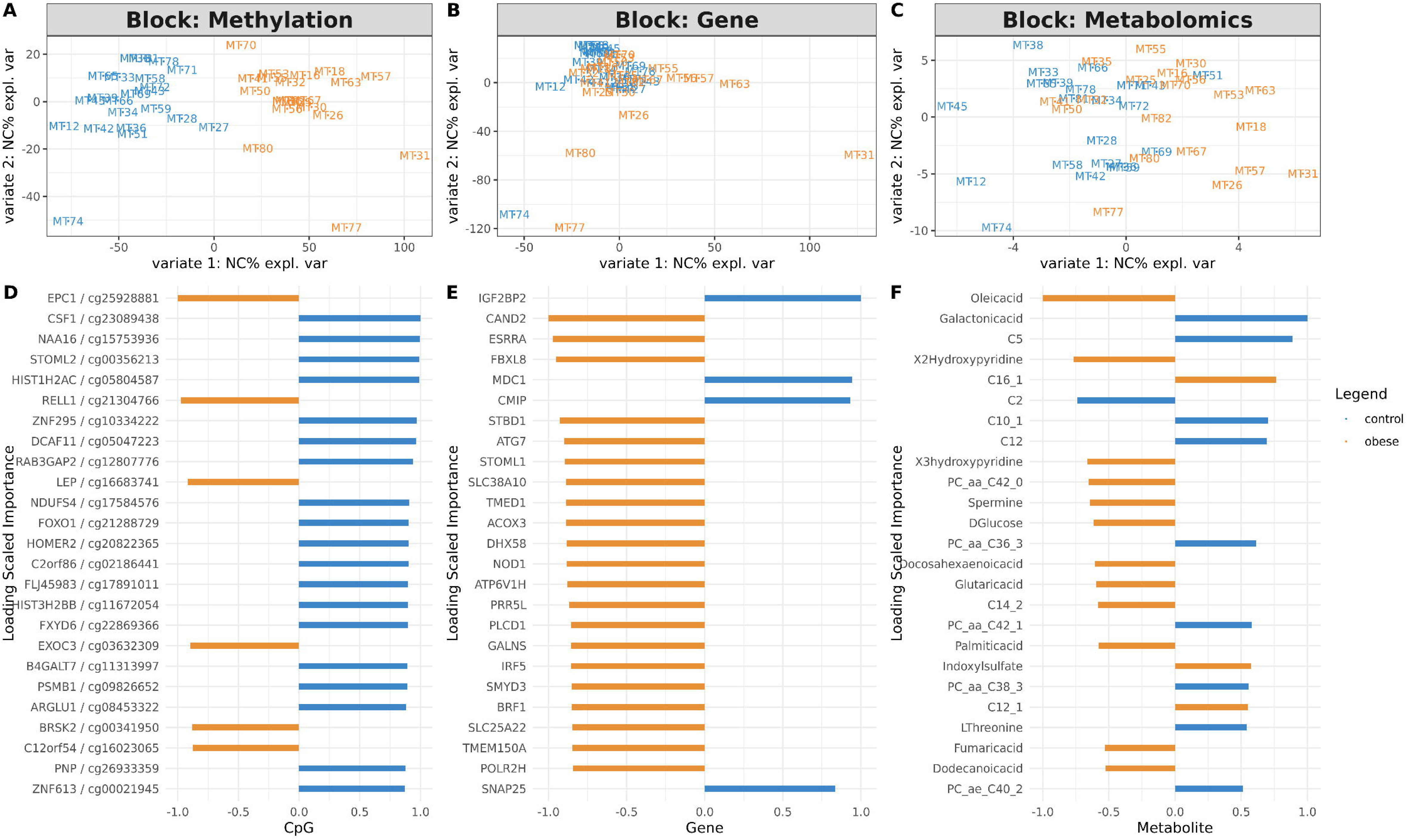
Multi-omics integration analysis. (**A-C**) Omics-specific sample plots from DIABLO showing the separation of obese and control samples in methylation data, gene expression data, and metabolomics data respectively. (**D-F**) Importance plot of top 25 features in methylation, gene expression and metabolomics modalities with the highest loadings extracted from the embedding space. The color represents the condition which features contribute the most.

The top 25 features from each omic with the highest feature weights (loadings) following integrated canonical correlation analysis are demonstrated in **Figure 5D-F**. The methylation features with the highest weights related to maternal obesity include CpG sites involved in cell-cycle control, glucose metabolism, and adipogenesis (FOXO1 ^91^), DNA repair (LIG3, SMUG1), erythropoietin pathway and differentiation (EPO, CSNK2A1, CSF1), which are hypermethylated in the obese group. Hypomethylation of LEP (encoding leptin) was also observed as a top feature, aligning with prior findings that maternal obesity is associated with elevated maternal leptin levels, a known marker of adipose tissue ^92^. These featured CpG sites indicate repression in fat metabolism and DNA repair and reduced differentiation potential. In the transcriptomic space, many genes related to mRNA splicing (SRRM1, IGF2BP1, IGF2BP2, CNOT4) have increased expression levels due to maternal obesity. Among the metabolite features, essential sugars (glucose, xylose), poly-unsaturated fatty acids (oleic acids, DHA, arachidonic acid), and phosphatidylcholine (PCs) are mostly decreased in the obese group; whereas most acylcarnitines (C) are elevated. The metabolic changes show an overall accumulation of saturated fatty acid, but repression of fat breakdown, glucose, and unsaturated fatty acid generation. As poly-unsaturated fatty acids (eg. arachidonic acid) have important anti-inflammatory effects, the results indicate a pro-inflammatory environment in offspring born of pre-pregnant obese mothers.

### The maternal obesity classification model is predictive of KIRC, LUSC, and PAAD cancers in TCGA

Maternal pre-pregnancy obesity may predispose a higher risk of cancer and other diseases in babies’ later life, via epigenetic modification ^12,17^. To check this assumption, we built maternal obesity random forest classification models using a total of 63 hypermethylated promoter region marker CpG sites obtained from top KEGG pathways which overlapped with the 14 TCGA cancer data that had sufficient numbers (n>10) of adjacent normal samples (**Supplemental Table 2**). The maternal obesity random forest model resulted in balanced accuracy of 0.93 on the obesity training data. We then applied this obesity classification model to predict the known adjacent normal and tumor tissue labels from DNA methylation data of 14 TCGA cancers, each of which has sufficient (n>10) tumor adjacent normal samples (**Figure 6**). This allows us to assess if the maternal obesity DNA methylation markers are associated with cancers. As shown in **Figure 6A**, three cancer types have good prediction balanced accuracy (Bal acc)of at least 0.7: LUSC (0.87), PAAD (0.83), KIRC (0.71), and two additional cancers reached 0.6: BRCA (0.60) and KIRP (0.63). The other metrics, including overall accuracy and F-1 scores are shown in **Figure 6B**. Thus, these results show that CpGs epigenetic markers associated with maternal obesity are also potentially associated with tumorigenesis in lung, breast, pancreas and kidney. Our result preliminarily supports that maternal pre-pregnancy obesity may predispose offspring to increased risks in certain cancers later in life through epigenetic modifications.

**Figure 6.**
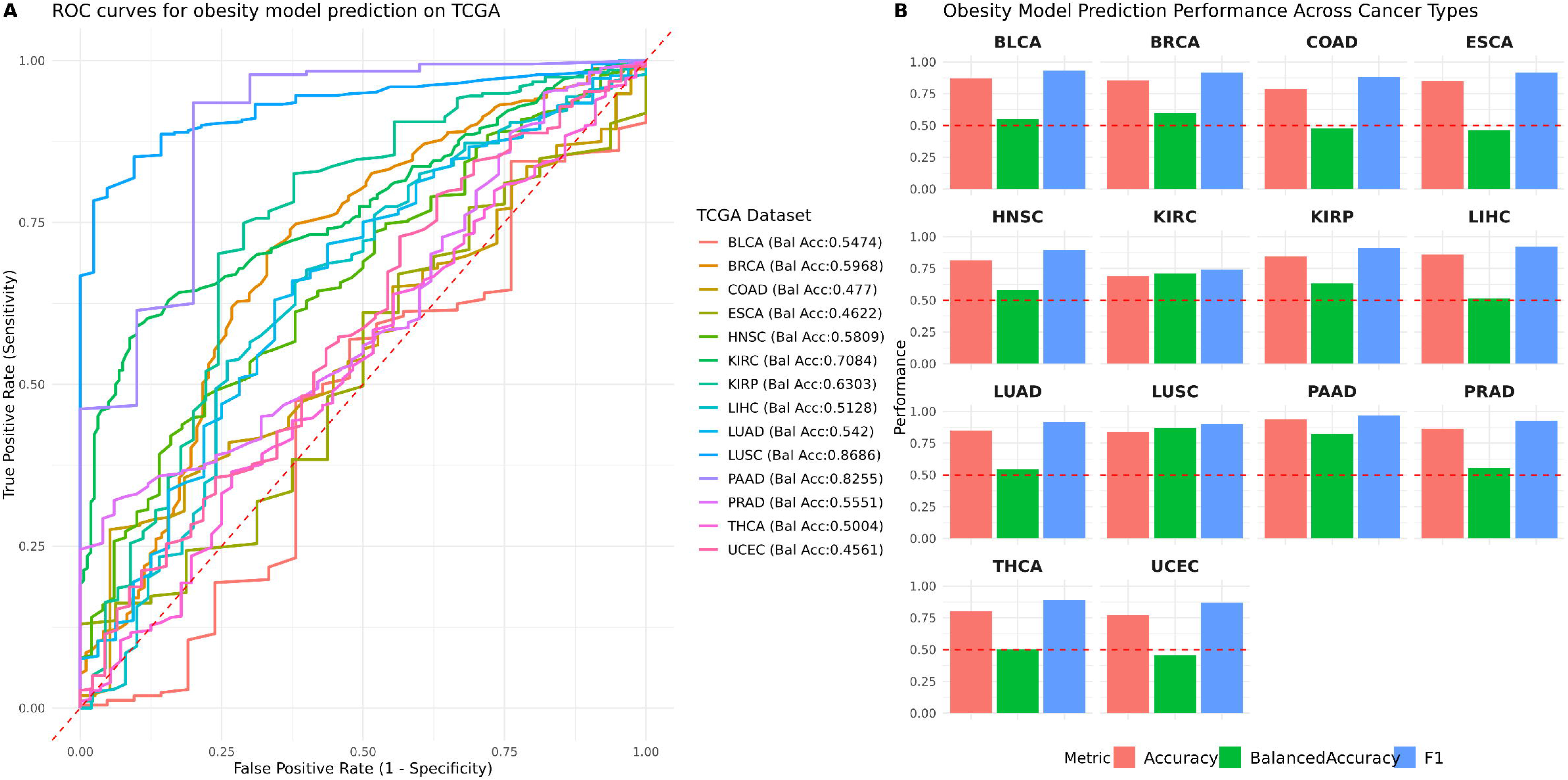
TCGA cancer classification by the maternal obesity classification model. (**A**) Receiver operating characteristic (ROC) curves on the 14 TCGA cancers, using the random forest classification model built with the CpG sites of genes in the top 5 pathways associated with maternal pre-pregnancy obesity. Balanced accuracy is shown for each cancer type. (**B**) Barplots showing the prediction performances on these TCGA datasets, using AUC, Balanced Accuracy, and F1 score.

## Discussion

Maternal obesity is one of the most urgent health concerns worldwide. Pre-pregnancy maternal obesity could cause various pregnancy-related complications and predispose offspring to cardiometabolic complications and chronic diseases in the long term ^9^. Multiple cross-continental large cohort meta-analyses have shown that maternal obesity is directly associated with offspring’s risk of obesity, coronary heart disease, insulin resistance, and adverse neurodevelopmental outcomes based on longitudinal observational studies ^9,93,94^. To directly pinpoint the molecular level changes in offspring by maternal pre-pregnancy obesity, we used cord blood stem cells as the studying material, which serve as a great surrogate revealing the newborn’s metabolism and immune system changes at the time of birth ^95^. The current study expands on previous effects and investigates the direct impact of maternal obesity on uHSCs programming, the progenitor of the immune cell population, using a multi-omics (epigenetic, gene expression, and metabolite) analysis approach from a unique multi-ethnic cohort.

Centered around methylation changes, three complimentary functional analysis approaches (KEGG, WGCNA, and PPI) consistently demonstrated that maternal obesity impacts multiple biological functions including hypermethylation in promoters of genes involved in cell cycle, ribosome biogenesis, and mTOR signaling pathways. Moreover, mTOR signaling pathway also plays a crucial role in metabolism and cell cycle regulation, disruption in this pathway leads to insulin resistance and long-term diseases ^96^. We observed a significant increase in stemness scores among uHSCs affected by maternal obesity, aligning with expected downregulation in the cell cycle gene expression due to observed hypermethylation in the promoters of these genes. Higher stemness scores indicate enhanced quiescence, shifting the balance between stem cell maintenance and differentiation towards the former. Unlike adult HSCs, fetal/neonatal HSCs typically exhibit higher proliferation and self-renewal capabilities, crucial for blood cell regeneration and innate immune system development ^97^. Our findings provide strong epigenetic evidence that maternal obesity compromises the maturation processes in neonatal uHSCs, which may predispose newborns to immunological disorders.

The subsequent multi-omics integration analysis expanded conclusions from methylation analysis to additional metabolomics readouts that are also linked to biological functions eg. cell cycle and inflammatory pathway. We thus propose the conceptual model to illustrate the effect of maternal pre-pregnancy obesity (**Figure 7**). Maternal obesity leads to nutrient deficiency with lower levels of essential amino acids and fatty acids in the newborn blood and disrupts the lipid metabolism homeosis in offspring. These metabolite changes further induce cell membrane instability and repress cell cycle progression, cell proliferation^98^, enhancing the dysregulation of these functions preexisting at the methylation level. Lipid dysregulation may also enhance the pro-inflammatory environment, which in turn induces complications in offspring later in life, such as cardiovascular diseases. Such a proposed model is also consistent with and further strengthens previous studies at the metabolomics or epigenome levels. For example, previous metabolomics studies of cord blood showed metabolic derangement predisposes newborns to cardiometabolic and endocrine diseases, and disrupt the normal hormone function and neonatal adiposity ^92,99^. Previous epigenome-wide association study (EWAS) with cord blood found a strong association between DNA methylation pattern and postnatal BMI trajectory until adolescent ^100^.

**Figure 7.**
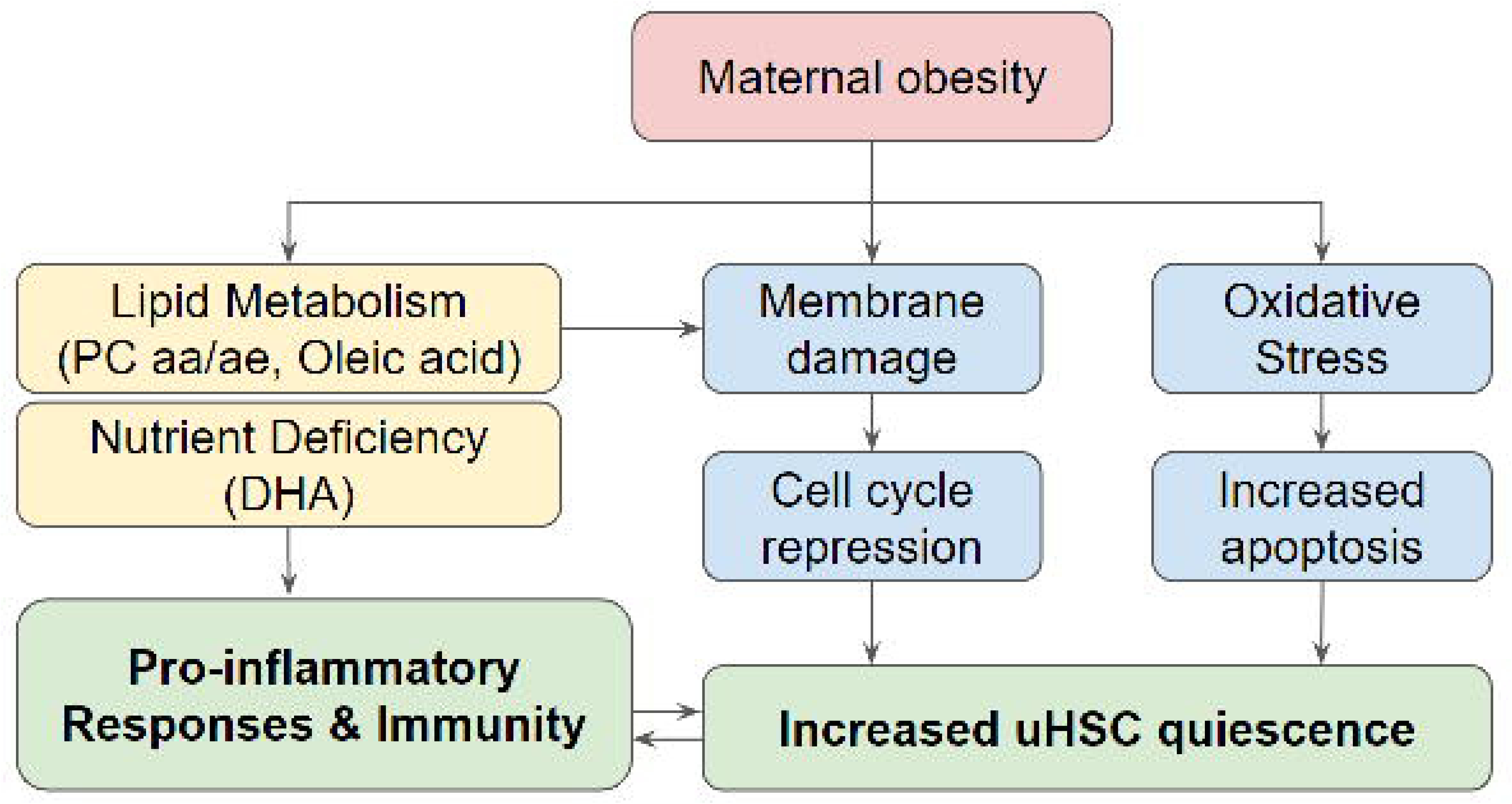
A proposed conceptual model of maternal obesity’s impact on neonatal development.

We also tested if maternal pre-pregnancy obesity can provide quantitative support to the long-speculated theory of the utero origin of cancers^10,11^. In particular, some researchers hypothesized there exist higher stem cell burdens in newborn babies born from obese mothers ^12^. Here we provide evidence that such stem cell burden is highly likely due to intergenerational DNA methylation modification on some key biological functions (cell cycle, ribosome function, and immune response) in the uHSCs. We built a random forest model trained on 61 maternal obesity-associated CpG markers in uHSCs and applied it to predict tumor and normal tissues across 14 TCGA cancer types, without prior cancer-specific training. This model achieved decent balanced accuracy above 0.6 for 5 out of 14 TCGA cancers investigated: LUSC, PAAD, BRCA KIRP and KIRC, reflecting its cross-context predictive potential. These cancers, characterized by inflammation, immune dysregulation, and epigenetic disruption, align with pathways enriched in the obesity-associated markers ^101,102^. Uncontrolled cell division, immune evasion, and chronic inflammation are well-established hallmarks of cancer ^103^, and these featured 61 CpG sites were implicated in relevant biological pathways that were intimately connected with cancer development. Aforehand results revealed a significant increase in stemness scores among uHSCs affected by maternal obesity, which aligns with the observed hypermethylation and subsequent downregulation of key cell cycle genes. This heightened stemness may predispose these cells to malignant transformation if these epigenetic modifications persist, leading to an elevated stem cell burden, disrupting normal cell cycle control, weakening immune surveillance, and ultimately increasing susceptibility to cancer. While performance was lower for other cancers, likely due to the small sample size limiting the detection of additional CpG biomarkers, tissue-specific methylation variability and microenvironment differences ^104^, the maternal obesity model implicates its biological relevance across diverse cancers.

This study is the first to directly examine the granular changes in the stem cell population of cord blood from babies of pre-pregnant obese mothers using a multi-omics approach. Previously, the association between maternal obesity and epigenetic modifications has been investigated across various tissue types (e.g., adipose tissue, liver, cord blood) and species (e.g., human, mouse) ^105^. We cross-checked our findings with these reports, many of which align providing further validation of their biological significance. For example, 33 CpGs across 20 genes, such as those in TAPBP (cg17621507, cg23922433, cg27385940), TNFAIP8 (cg18689486, cg07376834, cg03723497,cg21130861), and AGPAT1 (cg09043226, cg25733934, cg08049198, cg18191873) in our study are consistent with the cord blood leukocyte DNA methylation study from Martin et al with the same study objective ^106^. TAPBP, TNFAIP8, and AGPAT1 play roles in immune function, transcriptional regulation, and lipid metabolism respectively. Additionally, our analysis also identified different CpG probes within the same genes previously associated with maternal obesity, offering additional insights into their epigenetic regulation. For instance, we observed different CpGs in HLA-E (cg01462744, cg02942965, cg26175526), ALPK1 (cg04779144, cg10855342), and PTEN (cg09472211). These genes were also reported from the Boston Birth Cohort study ^107^. We identified different hypermethylation sites on MYT1L (cg05786278, cg17519749, cg21239227) and IGF1R (cg01284192, cg06596307, cg08138544, cg16918683, cg26577252), genes; these genes also showed high methylation levels in the cord blood (on different CpG sites) reported by Josefson et al ^108^. Additionally, in our gene expression and methylation correlation analysis of uHSC, HOXA9 and HOXA5 emerged as the top genes (**Supplemental Figure 3D**), displaying strong correlations between expression and methylation levels. These hypomethylated genes (HOX family genes), along with 25 additional commonly identified genes, are consistent with the finding in the DNA methylation study on leukocytes of cord blood samples ^109^, linking them to maternal lipid and cholesterol levels. In our study, HDAC4 and PLEC1 stand out for their strong associations with obesity-related traits among the top differentially methylated CpG sites. Hypermethylation of cg05995464 in HDAC4 was previously reported to be associated with childhood obesity ^110^. PLEC1 is a critical gene for extracellular matrix remodeling in adipose tissue, and hypomethylation of cg20784950 in PLEC1 is evident in our study. Lower PLEC1 methylation was previously correlated with higher BMI and obesity status ^83,111^. Together, these comparisons underscore the robust and overlapping epigenetic patterns associated with maternal pre-pregnancy obesity.

There are some caveats of this study related to the study design. First, this is a single-site study with a relatively small sample size, and along with some genomic inflation the statistical power of the findings is limited. This is especially the case for the gene expression omic layer, where individual DE genes are lacking. This may have limited maternal obesity CpG biomarker identification, which resulted in positive risk associations in some, but not all of the 14 TCGA cancers, in the classification model (**Figure 6**). When the budget allows, a large-scale multi-site population study is desirable. Secondly, we use the stem cell population in the cord blood as the surrogate for “stemness” property investigation, to link the offspring’s disease with maternal obesity. It is most feasible and practical to collect cord blood cells, and the painstaking measurements of the uHSC population avoid blood cell type heterogeneity issues, which may confound the cord blood DNA methylation result significantly ^81^. However, this approach may very well be simplified and biased, as stem cells exist in many body parts of babies. Therefore, extrapolations from uHSC need to be cautioned. Further, our phenotypic data collection focused on the physiological and demographic information and missed socioeconomic data. Thirdly, environmental, lifestyle or social determinants may act as confounders and influence the observed outcomes, which are not measured nor adjustable in the study, per the protocol. Some of these measurements, such as lifestyle and health insurance, can be mitigated by incorporating electronic health record data, similar to what we have done ^112,113^. Additionally, an important aspect of maternal-offspring study is to longitudinally follow them up for health outcomes later in life. The IRB for this study was not designed for such an investigation, unfortunately. Nevertheless, this uHSC multi-omics study provides a critical initial lens peeking into the immune-metabolic mechanisms, which serves as the foundation for all the possible expansion work mentioned above.

## Conclusion

In summary, this newborn study demonstrates the direct impact of maternal pre-pregnancy obesity and on newborn blood at the multi-omics level, which includes increased cell cycle arrest, impairment in the uHSCs differentiation capacity, more inflammation, and disruption in lipid metabolism. We also showed maternal obesity-associated epigenetic modifications are closely related to cancer markers, which could potentially help mitigate the intergenerational health risks.

## Disclosure of use of AI-assisted tools including generative AI

During the preparation of this work the author(s) used GPT-4.0 in order to improve the readability. Prompts used in GPT-4.0 include “help me improve my writing in a more logical and professional way” and “help me correct the grammar” along with a paragraph of the author’s own writing. After using this tool/service, the author(s) reviewed and edited the content thoroughly and take(s) full responsibility for the content of the publication.

## Data availability statement

DNA methylation data and bulk RNA-seq data generated in this study have been submitted and will be available through the National Institutes of Health Gene Expression Omnibus (GEO) with the accession number GSE273075 (GEO reviewer token: upmtkoygrlwtrmb). Other datasets used in this project for the analysis and validation purpose are publicly available. The placenta datasets used in this article are available in the GEO repository with accession numbers GSE31781, GSE36829, GSE59274, GSE44667, GSE74738, GSE49343, GSE69502, and GSE98224. Cord blood metabolomics data used in this article is available in metabolomics workbench with study ID ST001114. Cancer methylation datasets for BLCA, BRCA, COAD, ESCA, HNSC, KIRC, KIRP, LIHC, LUAD, LUSC, PAAD, PRAD, THCA, UCEC are available in The Cancer Genome Atlas (TCGA data portal: https://portal.gdc.cancer.gov/).

## Availability of source code and requirements

Project name: COBRE Hawaii Maternal Obesity Study

Project home page: https://github.com/lanagarmire/COBRE_methyl

Operating system(s): Windows, macOS, Linux

Programming language: R, Python

Other requirements: R ≥ 4.1.0

License: GNU-GPL-3.0

Code to produce the analyses in this manuscript are available through GitHub (https://github.com/lanagarmire/COBRE_methyl)

## Acknowledgments

This research was supported by grants R01 LM012373 and R01 LM012907 awarded by NLM, and R01 HD084633 awarded by NICHD to L.X. Garmire, as well as in part by the NCI Cancer Center Support Grant (CCSG) number P30 CA071789 awarded to Genomics and Bioinformatics Shared Resource (RRID:SCR_019085). This research was supported in part by training funding provided by the NIH grant T32 GM141746 and Advanced Proteogenomics of Cancer (T32 CA140044).

## Author contributions

LG envisioned this project, obtained the funding, supervised the study and revised the manuscript. YD performed the data analysis, generated the figures, and wrote the initial manuscript. YS collected TCGA data and built the machine learning models. RS consented patients and obtained the samples from the hospital, with coordination from PB. DT and SW coordinated with the patient recruitment and study. PB coordinated all the multi-omics assays. PB, CL, and FA designed the DNA methylation assays. AG performed FACS sorting of cord blood cells. ALJ performed the Illumina Meth 450 assay, MT supervised the Genomics Shared Resource analyses and provided a critical review of the manuscript. All authors have read the manuscript.

## Abbreviation

AA: Amino Acid
BH: Benjamini-Hochberg
BMI: Body mass index
C: Acylcarnitines
DE: Differential expression
DIABLO: Data Integration Analysis for Biomarker discovery using Latent cOmponents
DM: Differential methylation
DMR: Differentially methylated regions
DOHaD: Developmental Origins of Health and Disease
EWAS: Epigenome-wide association studies
FC: Fold Change
FDR: False positive results
KEGG: Kyoto Encyclopedia of Genes and Genomes
LOG: Logistic regression
MDS: Multi-dimensional Scaling
NHPI: Native Hawaiian and Pacific Islander
PANDA: Preferential Attachment-based common Neighbor Distribution derived Associations
PC aa: Diacyl phosphatidylcholines
PC ae: Acyl-alkylphosphatidylcholines
PCC: Pearson correlation coefficients
PPI: Protein-Protein Interaction
RF: Random Forest
SOV: Source of variance
SVD: Singular value decomposition
SVA: Surrogate variable analysis
TSS: Transcription start site
TCGA: The Cancer Genome Atlas
uHSCs: Umbilical cord blood hematopoietic stem cells
UMAP: Uniform Manifold Approximation and Projection
VSN: Variance Stabilization Normalization
WGCNA: Weighted Gene Co-expression Network Analysis

