## Supplementary Figures and Tables for "Multi-omics Analysis of Umbilical Cord Hematopoietic Stem Cells from a Multi-ethnic Cohort of Hawaii Reveals the Intergenerational Effect of Maternal Pre-Pregnancy Obesity and Risk Prediction for Cancers"

**Supplemental Figure 1. Data Processing Workflow.** The complete data pre-processing procedures consisted of sex mismatch check, filtration, quality control checks, normalization, batch correction, singular value decomposition analysis, and conversion of beta values to M-values. Created in https://BioRender.com.

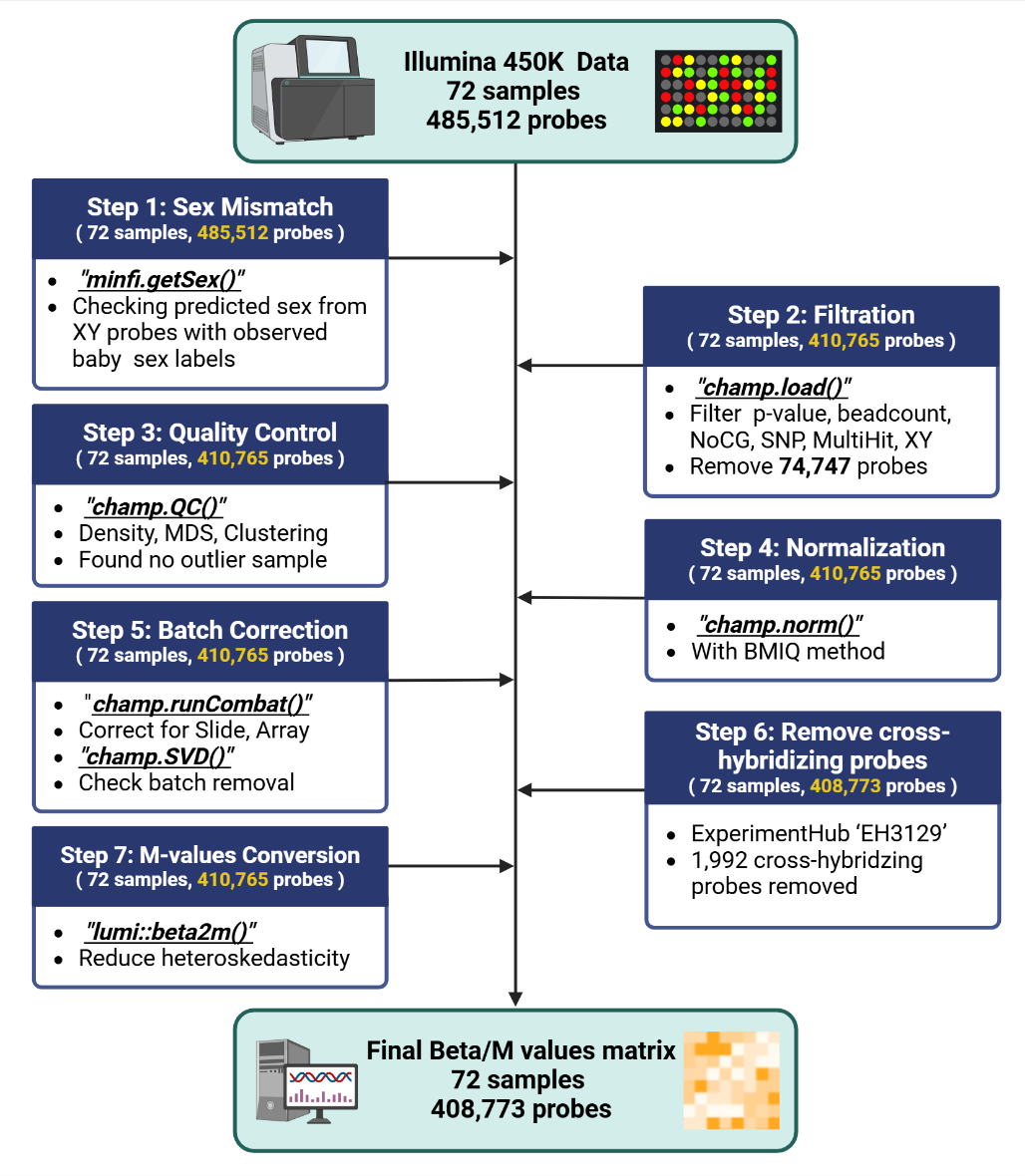

**Supplemental Figure 2. Data Quality Control.** (**A**) Sex mismatch check. (**B**) Median intensity plot. (**C**) MDS plot for top 1000 variable positions. (**D**) Raw density plot. (**E**) Singular value decomposition (SVD) plot after the removal of batch effects.

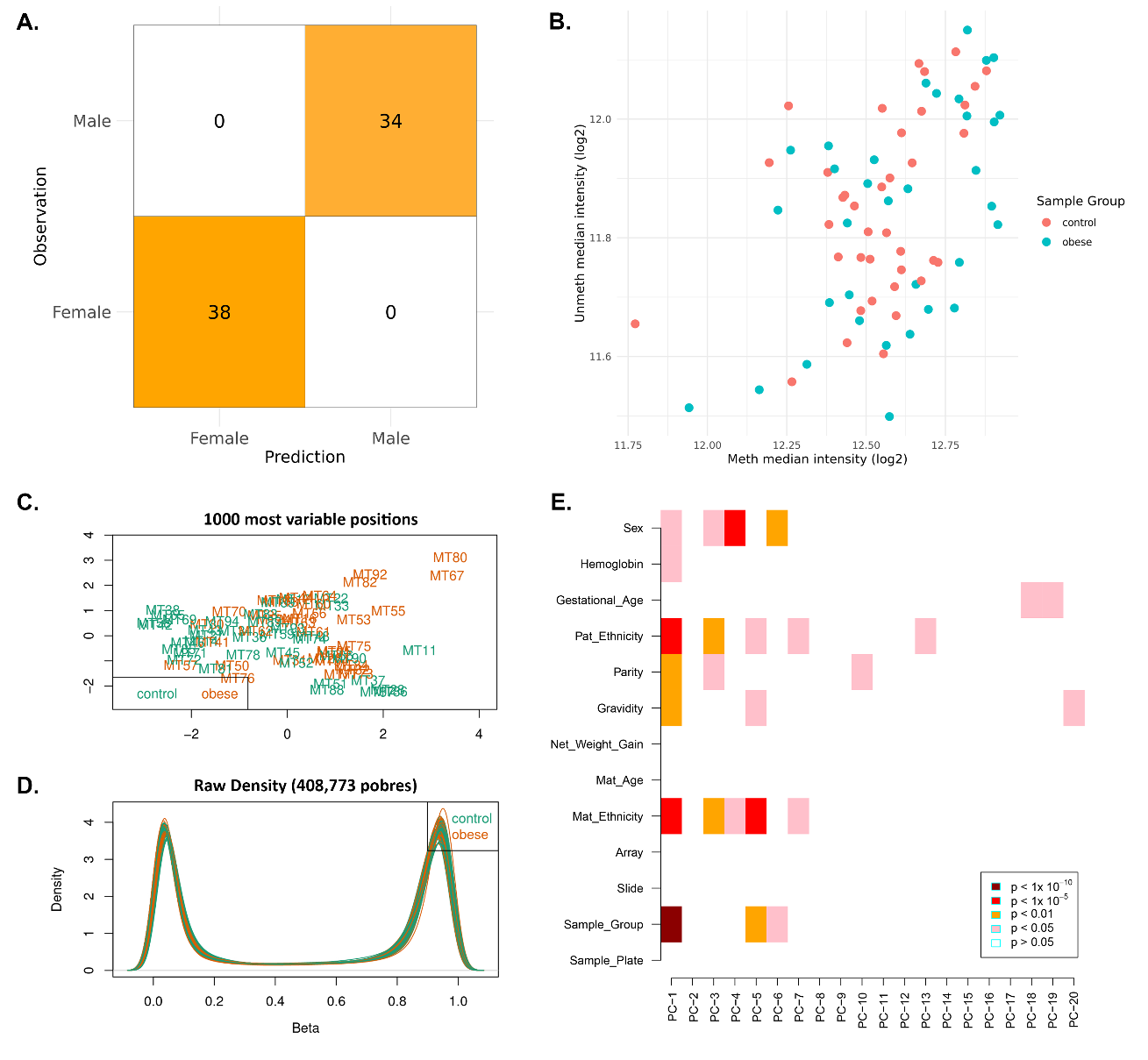

**Supplemental Figure 3. Differential expression and gene-methylation correlation analyses.**

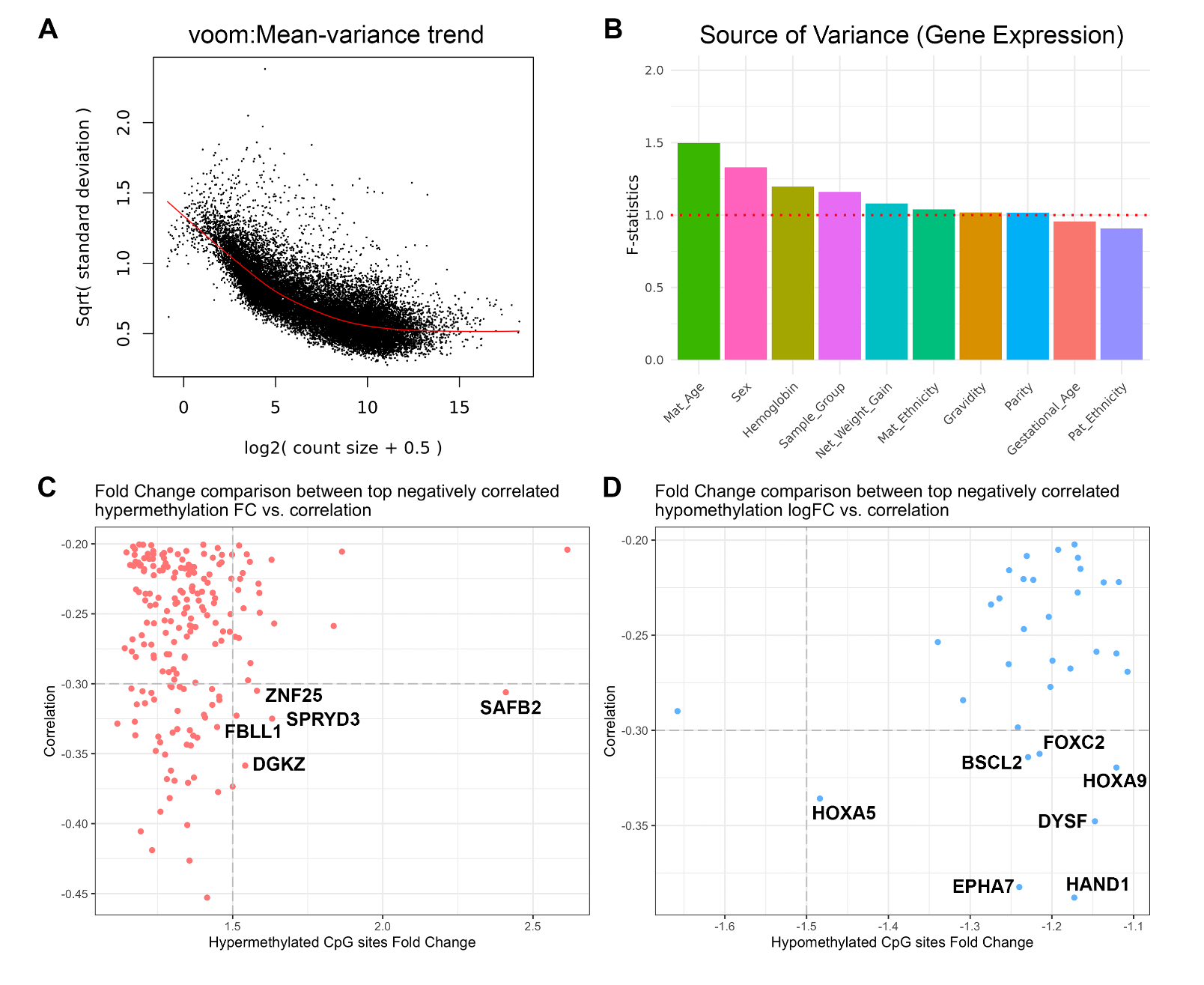

(A) limma-voom mean variance trend plot. (B) Source of variance analysis to build the differential expression confounder adjustment model. In the differential expression test, no significantly differentially expressed genes were found based on BH-adjusted p-values, so we checked the correlation between gene expression and DNA methylation levels for CpG sites from the promoter region (TSS200 and TSS1500) to find out if the different methylation signatures could be reflected on the gene level. (**C**) Scatter plot shows the correlation between methylation level and gene expression and the fold change of corresponding hypermethylated CpG sites between normal and obese patients. CpG sites of interest are annotated with gene symbols. (**D**) Scatter plot shows the correlation between methylation level and gene expression and the fold change of corresponding hypomethylated CpG sites between normal and obese patients. CpG sites of interest are annotated with gene symbols. Several genes are associated with cell apoptosis, adiposity, and metabolism with high negative Pearson correlations (PCC<-0.2) between bulk RNA-seq expression and methylation levels. Among these, SAFB2 stands out with a correlation of -0.306 and a significant fold change of 2.410, highlighting its critical roles in cell cycle control, differentiation, stress response, and the modulation of genes essential for apoptosis and immune functions^114^^,^^115^^,^^116^.

Furthermore, the functional enrichment of highly correlated genes with hypermethylated CpG sites reports multiple related pathways in **Supplementary Table 3**, including terpenoid backbone biosynthesis, endocrine resistance, and cell cycles. Genes related to cell proliferation and differentiation are also observed with high negative correlations to hypomethylated sites, including homeobox genes HOXA5 and HOXA9. These findings from the bulk RNA-seq analysis corroborate previous observations at the methylation level, suggesting that uHSCs from infants born to mothers with pre-pregnancy maternal obesity exhibit increased stemness due to alterations in cell cycle regulation.

**Supplemental Figure 4. Confounder adjustment Quantile-Quantile plot.** (**A**) Q-Q plot before confounder adjustment, only modeled with sample groups. (**B**) Q-Q plot after confounder adjustment. Genomic inflation scores before and after adjustment are reported as in lambda. The red line denotes the diagonal y=x line.

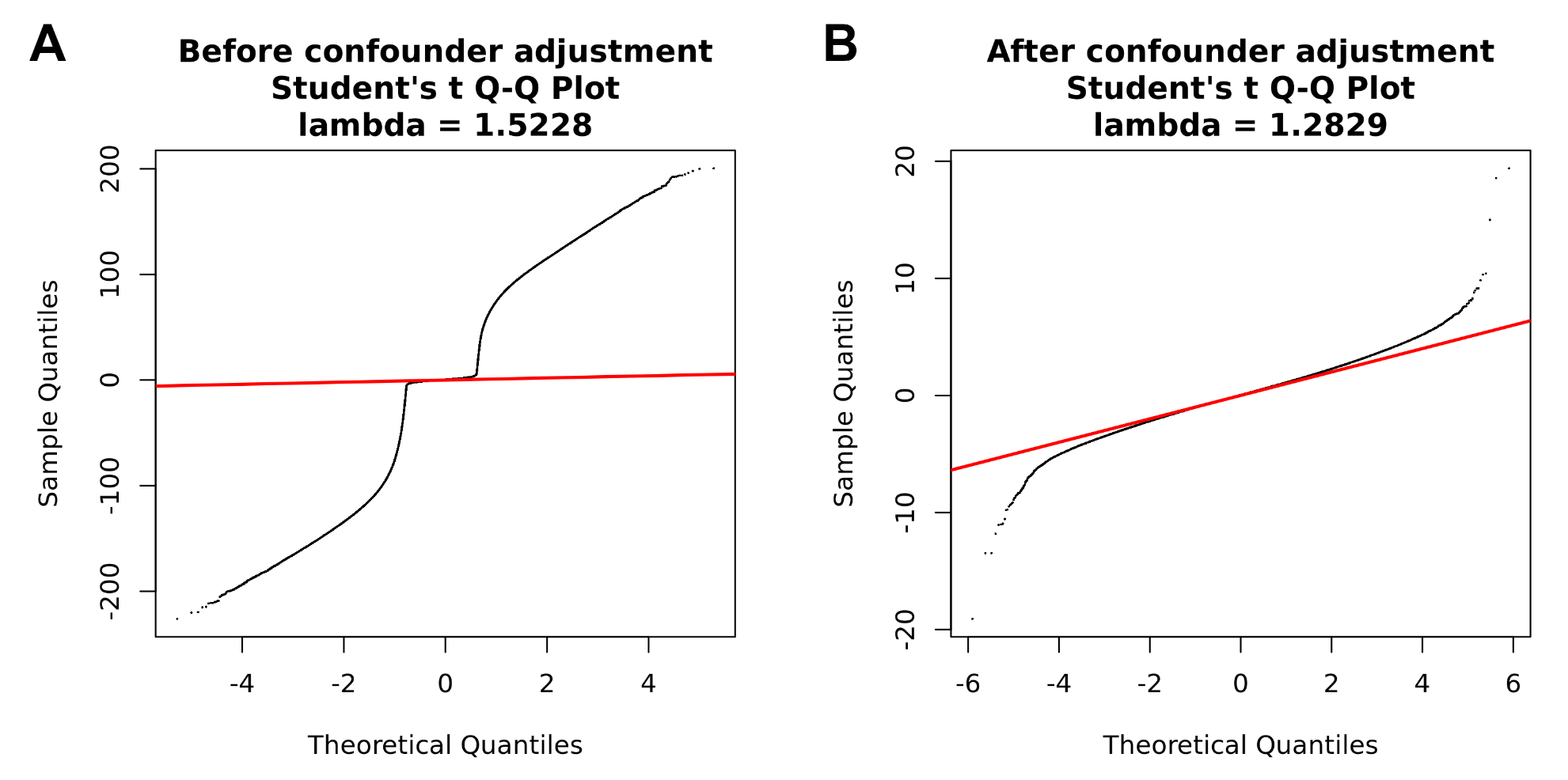

**Supplemental Figure 5. Validation on stem cell homogeneity.** UMAP plot of single cell profiles of two Barnyard controls (B1 and D1), non-maternally-obese control (C1, green), and maternally obese case (A1, red).

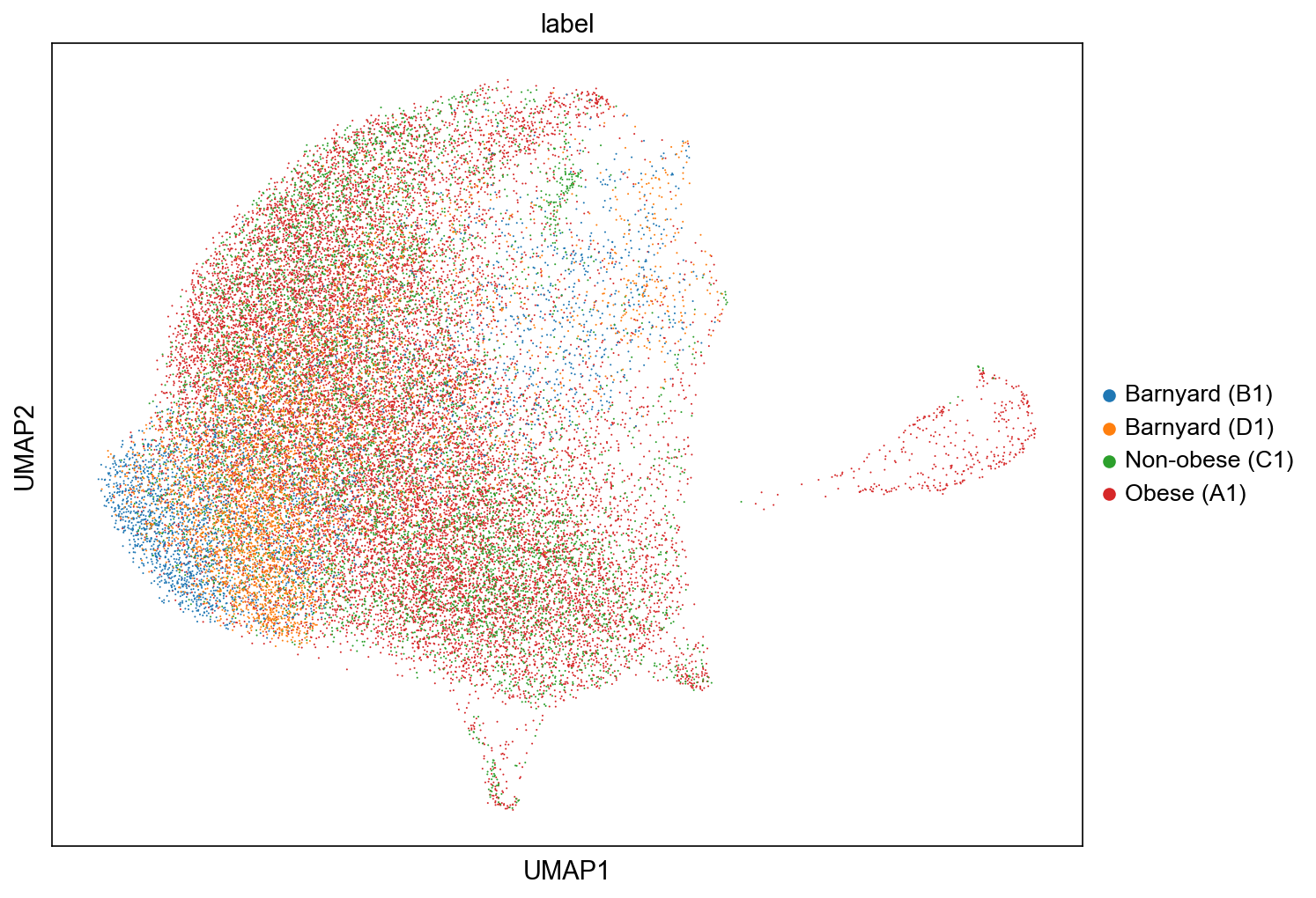

**Supplemental Figure 6. WGCNA modules.** (A) Module-trait association heatmap using beta values adjusted for confounding effects. (B) Complete gene networks including non-major WGCNA modules.

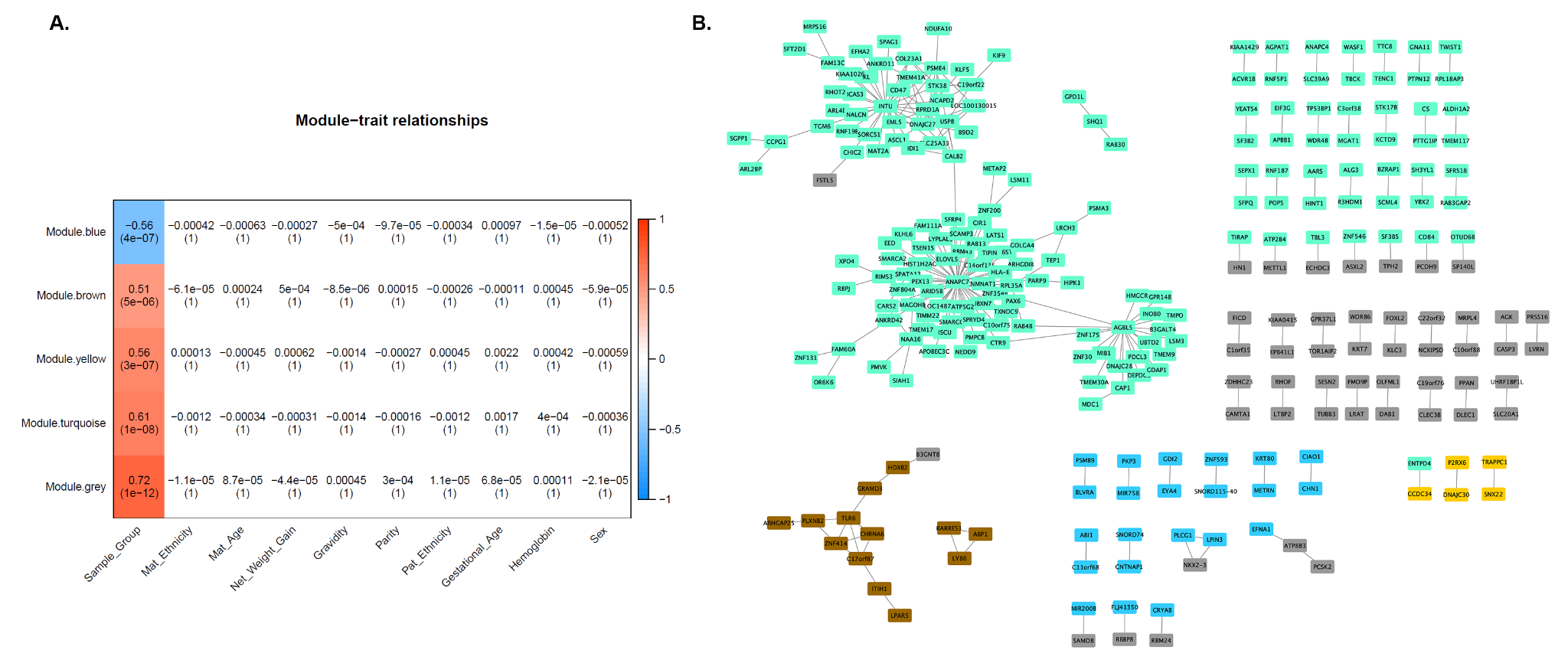

**Supplemental Table 1. Gestational age associated with differentially methylated regions.**

| seqnames | start | end | width | value | area | p.value | fwer | p.valueArea | GEOAC |
| --- | --- | --- | --- | --- | --- | --- | --- | --- | --- |
| chr16 | 30490382 | 30490557 | 175 | 0.00753 | 0.052707 | 0 | 0 | 0 | GSE31781 |
| chr16 | 66614096 | 66614713 | 617 | 0.007503 | 0.052518 | 0 | 0 | 0 | GSE31781 |
| chr3 | 45705493 | 45706258 | 765 | 0.005333 | 0.047996 | 0 | 0 | 0 | GSE31781 |
| chr19 | 10689819 | 10690176 | 357 | 0.00518 | 0.036259 | 0 | 0 | 0 | GSE31781 |
| chr12 | 15366503 | 15367295 | 792 | -0.00571 | 0.034242 | 0 | 0 | 0 | GSE31781 |
| chr3 | 61211587 | 61212310 | 723 | 0.004155 | 0.033238 | 0 | 0 | 0 | GSE31781 |
| chr12 | 4253246 | 4254283 | 1037 | 0.003561 | 0.032048 | 0 | 0 | 0 | GSE31781 |
| chr19 | 14501235 | 14501403 | 168 | 0.005077 | 0.030464 | 0 | 0 | 0 | GSE31781 |
| chr10 | 131155063 | 131155565 | 502 | 0.004013 | 0.016051 | 0 | 0 | 0.006678 | GSE31781 |
| chr11 | 14949954 | 14950394 | 440 | 0.002857 | 0.014287 | 0 | 0 | 0.011686 | GSE31781 |
| chr19 | 14501456 | 14501546 | 90 | 0.005063 | 0.010126 | 0 | 0 | 0.016694 | GSE31781 |
| chr9 | 92603597 | 92603970 | 373 | 0.004005 | 0.00801 | 0 | 0 | 0.025042 | GSE31781 |
| chr12 | 15366586 | 15366586 | 0 | -0.00753 | 0.007528 | 0 | 0 | 0.033389 | GSE31781 |
| chr9 | 92603599 | 92603599 | 0 | 0.005069 | 0.005069 | 0 | 0 | 0.113523 | GSE31781 |
| chr9 | 92603849 | 92603849 | 0 | 0.003609 | 0.003609 | 0.001669 | 0.001 | 0.268781 | GSE31781 |
| chr15 | 31195708 | 31196075 | 367 | -0.02866 | 0.229244 | 0.001143 | 0.243 | 0.029675 | GSE74738 |
| chr1 | 161008127 | 161008977 | 850 | 0.024888 | 0.248883 | 0.002302 | 0.425 | 0.024101 | GSE74738 |
| chr5 | 139493486 | 139493680 | 194 | -0.0357 | 0.321332 | 0.004982 | 0.629 | 0.011308 | GSE74738 |
| chr3 | 43020564 | 43021399 | 835 | -0.03606 | 0.288508 | 0.00564 | 0.659 | 0.015818 | GSE74738 |
| chr1 | 6684860 | 6685162 | 302 | 0.023993 | 0.167953 | 0.010519 | 0.876 | 0.053484 | GSE74738 |
| chr6 | 28945182 | 28945507 | 325 | -0.02712 | 0.216945 | 0.014034 | 0.904 | 0.033489 | GSE74738 |
| chr16 | 3508542 | 3508542 | 0 | 0.042375 | 0.042375 | 0.014737 | 0.792 | 0.278253 | GSE74738 |
| chr19 | 58220295 | 58220837 | 542 | -0.02055 | 0.164411 | 0.017672 | 0.941 | 0.055181 | GSE74738 |
| chr3 | 45883249 | 45883859 | 610 | -0.02405 | 0.26456 | 0.019916 | 0.949 | 0.020487 | GSE74738 |
| chr2 | 70995349 | 70995607 | 258 | 0.031195 | 0.155975 | 0.021881 | 0.951 | 0.059164 | GSE74738 |
| chr16 | 3508257 | 3508546 | 289 | 0.033495 | 0.06699 | 0.022789 | 0.941 | 0.151711 | GSE74738 |
| chr7 | 11871535 | 11872073 | 538 | -0.02785 | 0.222823 | 0.023883 | 0.954 | 0.031582 | GSE74738 |
| chr6 | 28411030 | 28411423 | 393 | -0.0264 | 0.184813 | 0.026103 | 0.961 | 0.045801 | GSE74738 |
| chr3 | 148804272 | 148804272 | 0 | -0.0382 | 0.038202 | 0.027311 | 0.918 | 0.332292 | GSE74738 |
| chr2 | 70995440 | 70995522 | 82 | 0.031306 | 0.062612 | 0.030793 | 0.965 | 0.166452 | GSE74738 |
| chr3 | 148804275 | 148804275 | 0 | -0.03736 | 0.03736 | 0.030928 | 0.932 | 0.338742 | GSE74738 |
| chr10 | 99790170 | 99790205 | 35 | -0.03197 | 0.06393 | 0.031257 | 0.96 | 0.161832 | GSE74738 |
| chr7 | 101005910 | 101006573 | 663 | -0.02655 | 0.238909 | 0.032906 | 0.977 | 0.026884 | GSE74738 |
| chr4 | 93226245 | 93227195 | 950 | -0.02759 | 0.110356 | 0.039105 | 0.989 | 0.084761 | GSE74738 |
| chr2 | 30669597 | 30669597 | 0 | 0.062872 | 0.062872 | 0.000205 | 0.037 | 0.093345 | GSE69502 |
| chr12 | 29301525 | 29302714 | 1189 | -0.02929 | 0.263594 | 0.000814 | 0.183 | 0.004139 | GSE69502 |
| chr10 | 42862876 | 42863594 | 718 | -0.01808 | 0.144628 | 0.001814 | 0.317 | 0.027638 | GSE69502 |
| chr2 | 30669759 | 30669759 | 0 | 0.035586 | 0.035586 | 0.003852 | 0.438 | 0.189043 | GSE69502 |
| chr19 | 9785295 | 9786077 | 782 | -0.02752 | 0.275236 | 0.003879 | 0.484 | 0.003472 | GSE69502 |
| chr2 | 70994802 | 70995607 | 805 | -0.01909 | 0.229087 | 0.006053 | 0.695 | 0.007154 | GSE69502 |
| chr5 | 67583972 | 67584380 | 408 | 0.016011 | 0.128091 | 0.010467 | 0.846 | 0.035938 | GSE69502 |
| chr8 | 67344553 | 67345006 | 453 | -0.01443 | 0.129889 | 0.016287 | 0.918 | 0.034935 | GSE69502 |
| chr17 | 3438933 | 3439042 | 109 | 0.020569 | 0.041137 | 0.024991 | 0.91 | 0.157832 | GSE69502 |
| chr7 | 4784887 | 4784903 | 16 | -0.02107 | 0.042145 | 0.027549 | 0.905 | 0.152682 | GSE69502 |
| chr17 | 36105064 | 36105517 | 453 | 0.014932 | 0.104523 | 0.031435 | 0.956 | 0.050598 | GSE69502 |
| chr2 | 20101506 | 20101506 | 0 | 0.024595 | 0.024595 | 0.031625 | 0.896 | 0.316807 | GSE69502 |
| chr12 | 53693322 | 53693485 | 163 | -0.01599 | 0.079944 | 0.040065 | 0.974 | 0.070606 | GSE69502 |
| chr17 | 3438918 | 3438918 | 0 | 0.023404 | 0.023404 | 0.041534 | 0.926 | 0.329692 | GSE69502 |
| chr8 | 26047780 | 26048281 | 501 | 0.013101 | 0.078605 | 0.041825 | 0.98 | 0.072059 | GSE69502 |
| chr4 | 109092628 | 109092628 | 0 | 0.023172 | 0.023172 | 0.0437 | 0.929 | 0.33087 | GSE69502 |
| chr1 | 86622113 | 86622737 | 624 | 0.013974 | 0.097819 | 0.045386 | 0.983 | 0.055411 | GSE69502 |
| chr3 | 44667010 | 44667010 | 0 | 0.023006 | 0.023006 | 0.045502 | 0.934 | 0.331874 | GSE69502 |
| chr17 | 3438857 | 3439203 | 346 | 0.019457 | 0.038913 | 0.045797 | 0.951 | 0.170589 | GSE69502 |
| chr1 | 47488757 | 47488757 | 0 | 0.022784 | 0.022784 | 0.047866 | 0.938 | 0.333141 | GSE69502 |
| chr2 | 176947764 | 176949017 | 1253 | -0.00765 | 0.107098 | 0.000927 | 0.223 | 0.01778 | GSE98224 |
| chr15 | 31195612 | 31196075 | 463 | -0.00742 | 0.066777 | 0.002565 | 0.558 | 0.05018 | GSE98224 |
| chr2 | 214148856 | 214149425 | 569 | -0.01433 | 0.14331 | 0.00283 | 0.487 | 0.007047 | GSE98224 |
| chr2 | 70994802 | 70995607 | 805 | -0.01467 | 0.176081 | 0.003151 | 0.515 | 0.003092 | GSE98224 |
| chr8 | 125985352 | 125985883 | 531 | -0.01545 | 0.108126 | 0.003477 | 0.523 | 0.017282 | GSE98224 |
| chr14 | 95234658 | 95235489 | 831 | 0.015054 | 0.135485 | 0.004254 | 0.567 | 0.008651 | GSE98224 |
| chr5 | 139493486 | 139493680 | 194 | 0.014412 | 0.129707 | 0.004258 | 0.604 | 0.010052 | GSE98224 |
| chr17 | 77923971 | 77924733 | 762 | 0.01443 | 0.115437 | 0.004987 | 0.623 | 0.014449 | GSE98224 |
| chr8 | 15397637 | 15398333 | 696 | 0.013362 | 0.106898 | 0.005892 | 0.698 | 0.017858 | GSE98224 |
| chr7 | 96650668 | 96650668 | 0 | 0.020293 | 0.020293 | 0.006135 | 0.666 | 0.20312 | GSE98224 |
| chr1 | 6684860 | 6685162 | 302 | 0.010645 | 0.074512 | 0.007122 | 0.768 | 0.041552 | GSE98224 |
| chr10 | 8094641 | 8094860 | 219 | 0.016152 | 0.032304 | 0.007548 | 0.648 | 0.1151 | GSE98224 |
| chr6 | 33041218 | 33041268 | 50 | 0.01471 | 0.04413 | 0.009066 | 0.719 | 0.082296 | GSE98224 |
| chr10 | 13749010 | 13749010 | 0 | 0.018125 | 0.018125 | 0.010064 | 0.755 | 0.22644 | GSE98224 |
| chr2 | 223177596 | 223177596 | 0 | -0.01686 | 0.016855 | 0.013985 | 0.813 | 0.247797 | GSE98224 |
| chr18 | 42530797 | 42530935 | 138 | 0.012265 | 0.036794 | 0.014617 | 0.864 | 0.098827 | GSE98224 |
| chr5 | 114937614 | 114937919 | 305 | 0.013743 | 0.027486 | 0.015312 | 0.797 | 0.140932 | GSE98224 |
| chr18 | 42530692 | 42530893 | 201 | 0.014173 | 0.028346 | 0.015432 | 0.788 | 0.135665 | GSE98224 |
| chr16 | 81039227 | 81039227 | 0 | 0.016417 | 0.016417 | 0.015817 | 0.835 | 0.257622 | GSE98224 |
| chr11 | 74302893 | 74302893 | 0 | 0.01641 | 0.01641 | 0.015839 | 0.835 | 0.257752 | GSE98224 |
| chr11 | 111250093 | 111250566 | 473 | 0.009577 | 0.095774 | 0.015922 | 0.922 | 0.02394 | GSE98224 |
| chr10 | 8094482 | 8094679 | 197 | 0.013857 | 0.027713 | 0.01812 | 0.816 | 0.139474 | GSE98224 |
| chr6 | 33240864 | 33241410 | 546 | 0.011451 | 0.045803 | 0.018875 | 0.904 | 0.079444 | GSE98224 |
| chr10 | 102279373 | 102279694 | 321 | -0.01263 | 0.050531 | 0.019462 | 0.868 | 0.072049 | GSE98224 |
| chr5 | 114938002 | 114938002 | 0 | 0.015692 | 0.015692 | 0.019836 | 0.879 | 0.276512 | GSE98224 |
| chr5 | 126409227 | 126409227 | 0 | 0.01565 | 0.01565 | 0.020086 | 0.879 | 0.277659 | GSE98224 |
| chr6 | 88875236 | 88875236 | 0 | -0.01561 | 0.015615 | 0.020299 | 0.88 | 0.278665 | GSE98224 |
| chr8 | 21916256 | 21917015 | 759 | 0.008803 | 0.061619 | 0.022109 | 0.944 | 0.056745 | GSE98224 |
| chr6 | 32060681 | 32061478 | 797 | 0.008349 | 0.06679 | 0.0224 | 0.95 | 0.050161 | GSE98224 |
| chr5 | 142784982 | 142785258 | 276 | -0.01294 | 0.025875 | 0.022767 | 0.865 | 0.150226 | GSE98224 |
| chr19 | 1275266 | 1275762 | 496 | -0.01134 | 0.045377 | 0.022897 | 0.92 | 0.080251 | GSE98224 |
| chr5 | 44388688 | 44389406 | 718 | -0.0109 | 0.043591 | 0.023073 | 0.925 | 0.083253 | GSE98224 |
| chr13 | 88324193 | 88324879 | 686 | -0.01116 | 0.066983 | 0.023649 | 0.927 | 0.049903 | GSE98224 |
| chr7 | 114055123 | 114055204 | 81 | 0.01195 | 0.047799 | 0.024094 | 0.907 | 0.076304 | GSE98224 |
| chr12 | 51717674 | 51718355 | 681 | 0.010245 | 0.112692 | 0.024168 | 0.941 | 0.015424 | GSE98224 |
| chr10 | 102279330 | 102279703 | 373 | -0.0121 | 0.036306 | 0.024954 | 0.904 | 0.10039 | GSE98224 |
| chr6 | 33041229 | 33041343 | 114 | 0.011392 | 0.022785 | 0.025817 | 0.927 | 0.175725 | GSE98224 |
| chr1 | 35220334 | 35220710 | 376 | -0.00985 | 0.039385 | 0.026 | 0.949 | 0.092053 | GSE98224 |
| chr7 | 114055074 | 114055210 | 136 | 0.012567 | 0.025133 | 0.026236 | 0.888 | 0.155086 | GSE98224 |
| chr1 | 50573970 | 50573970 | 0 | 0.014791 | 0.014791 | 0.026408 | 0.897 | 0.305211 | GSE98224 |
| chr11 | 124767720 | 124767974 | 254 | 0.010905 | 0.043619 | 0.026527 | 0.936 | 0.083197 | GSE98224 |
| chr8 | 144965577 | 144965915 | 338 | 0.011378 | 0.034133 | 0.026531 | 0.928 | 0.108158 | GSE98224 |
| chr11 | 124767840 | 124768015 | 175 | 0.011206 | 0.022412 | 0.027589 | 0.934 | 0.179875 | GSE98224 |
| chr7 | 96650192 | 96650509 | 317 | 0.011023 | 0.03307 | 0.02763 | 0.94 | 0.111994 | GSE98224 |
| chr5 | 114937535 | 114938236 | 701 | 0.011853 | 0.047411 | 0.027888 | 0.918 | 0.076823 | GSE98224 |
| chr10 | 71812265 | 71812630 | 365 | -0.00995 | 0.069631 | 0.028228 | 0.946 | 0.046767 | GSE98224 |
| chr6 | 30181877 | 30182240 | 363 | -0.01243 | 0.024852 | 0.031066 | 0.908 | 0.157149 | GSE98224 |
| chr2 | 198650752 | 198651576 | 824 | -0.00978 | 0.048881 | 0.032109 | 0.953 | 0.074562 | GSE98224 |
| chr5 | 126409211 | 126409211 | 0 | 0.014204 | 0.014204 | 0.032288 | 0.915 | 0.326998 | GSE98224 |
| chr2 | 223177574 | 223177574 | 0 | -0.01414 | 0.014138 | 0.033013 | 0.917 | 0.329551 | GSE98224 |
| chr12 | 117036703 | 117036972 | 269 | 0.00927 | 0.037078 | 0.033765 | 0.966 | 0.098087 | GSE98224 |
| chr5 | 153853057 | 153853545 | 488 | -0.00996 | 0.039827 | 0.033929 | 0.959 | 0.091081 | GSE98224 |
| chr10 | 2931961 | 2932386 | 425 | 0.007398 | 0.02959 | 0.034142 | 0.976 | 0.128641 | GSE98224 |
| chr2 | 42067938 | 42068648 | 710 | -0.00737 | 0.058975 | 0.03427 | 0.976 | 0.059978 | GSE98224 |
| chr7 | 96626399 | 96627050 | 651 | 0.010033 | 0.0301 | 0.035559 | 0.961 | 0.125923 | GSE98224 |
| chr11 | 78387190 | 78387503 | 313 | 0.009323 | 0.055939 | 0.036961 | 0.965 | 0.064064 | GSE98224 |
| chr6 | 30181713 | 30181771 | 58 | -0.01164 | 0.023272 | 0.03867 | 0.932 | 0.170723 | GSE98224 |
| chr11 | 105481509 | 105481509 | 0 | -0.01366 | 0.013658 | 0.039185 | 0.94 | 0.346841 | GSE98224 |
| chr4 | 165304842 | 165305050 | 208 | -0.01022 | 0.030664 | 0.039477 | 0.965 | 0.122828 | GSE98224 |
| chr5 | 142784721 | 142785172 | 451 | -0.01126 | 0.022525 | 0.039795 | 0.942 | 0.17857 | GSE98224 |
| chr2 | 134326366 | 134326366 | 0 | 0.013609 | 0.013609 | 0.039851 | 0.94 | 0.348624 | GSE98224 |
| chr1 | 236686598 | 236687575 | 977 | -0.00839 | 0.067142 | 0.039873 | 0.975 | 0.049716 | GSE98224 |
| chr6 | 32116905 | 32116994 | 89 | -0.01117 | 0.022346 | 0.041769 | 0.943 | 0.180596 | GSE98224 |
| chr6 | 33868187 | 33868466 | 279 | 0.008484 | 0.025451 | 0.0435 | 0.976 | 0.152936 | GSE98224 |
| chr6 | 32116653 | 32116963 | 310 | -0.01082 | 0.043287 | 0.04369 | 0.958 | 0.083773 | GSE98224 |
| chr6 | 28411030 | 28411378 | 348 | -0.00822 | 0.049318 | 0.043877 | 0.979 | 0.073889 | GSE98224 |
| chr7 | 99516845 | 99517279 | 434 | -0.01 | 0.040018 | 0.044995 | 0.97 | 0.0907 | GSE98224 |
| chr6 | 31543540 | 31543686 | 146 | -0.00791 | 0.047436 | 0.045021 | 0.98 | 0.076805 | GSE98224 |
| chr6 | 30181645 | 30181936 | 291 | -0.0113 | 0.033904 | 0.045959 | 0.947 | 0.108846 | GSE98224 |
| chr10 | 8094534 | 8094534 | 0 | 0.013178 | 0.013178 | 0.046602 | 0.95 | 0.358931 | GSE98224 |
| chr17 | 48172203 | 48172203 | 0 | -0.01308 | 0.013085 | 0.048322 | 0.95 | 0.359705 | GSE98224 |
| chr6 | 30181942 | 30181942 | 0 | -0.01307 | 0.013068 | 0.048602 | 0.951 | 0.359877 | GSE98224 |
| chr6 | 32027623 | 32027666 | 43 | 0.010806 | 0.021612 | 0.04907 | 0.96 | 0.188671 | GSE98224 |
| chr5 | 67584194 | 67584380 | 186 | 0.007055 | 0.035274 | 0.049137 | 0.987 | 0.103971 | GSE98224 |
| chr10 | 86001154 | 86001560 | 406 | 0.008535 | 0.059745 | 0.049784 | 0.977 | 0.058958 | GSE98224 |

Gestational age associated with differentially methylated regions (DMRs) were reported from public pregnancy-related methylation datasets. Starting and ending positions of the DMRs were listed with p.value < 0.05.

**Supplemental Table 2. List of CpG feature used in maternal obesity classification model for TCGA cancer prediction.** CpG features that are used for maternal obesity classification model building, gene annotation, promotor regions, and related KEGG pathways are listed respectively.

| CpG | Gene | Group | Pathway |
| --- | --- | --- | --- |
| cg00174179 | RHOA | TSS1500 | mTOR signaling pathway |
| cg00199549 | MCM5 | TSS1500 | Cell cycle |
| cg00468144 | WNT2B | TSS1500 | mTOR signaling pathway |
| cg01051310 | WNT3A | TSS1500 | mTOR signaling pathway |
| cg01299496 | NOP10 | TSS200 | Ribosome biogenesis in eukaryotes |
| cg01435220 | FZD2 | TSS1500 | mTOR signaling pathway |
| cg01445659 | MRPL34 | TSS1500 | Ribosome |
| cg02017282 | WNT3 | TSS1500 | mTOR signaling pathway |
| cg02789441 | IMP4 | TSS200 | Ribosome biogenesis in eukaryotes |
| cg02929734 | E2F4 | TSS200 | Cell cycle |
| cg02951971 | SOS2 | TSS1500 | mTOR signaling pathway |
| cg03074424 | NUP35 | TSS200 | Nucleocytoplasmic transport |
| cg03366382 | INS | TSS1500 | mTOR signaling pathway |
| cg03471150 | IPO9 | TSS1500 | Nucleocytoplasmic transport |
| cg03617406 | SMAD2 | TSS200 | Cell cycle |
| cg04025728 | RPL22 | TSS200 | Ribosome |
| cg04798158 | FCF1 | TSS200 | Ribosome biogenesis in eukaryotes |
| cg05569124 | CHEK1 | TSS1500 | Cell cycle |
| cg05925507 | NXF1 | TSS200 | Nucleocytoplasmic transport, Ribosome biogenesis in eukaryotes |
| cg06143615 | NUP210 | TSS1500 | Nucleocytoplasmic transport |
| cg06513075 | NAT10 | TSS1500 | Ribosome biogenesis in eukaryotes |
| cg06536629 | MCM6 | TSS200 | Cell cycle |
| cg06710522 | MRPL10 | TSS200 | Ribosome |
| cg07087018 | NUP93 | TSS200 | Nucleocytoplasmic transport |
| cg07896667 | WNT5B | TSS200 | mTOR signaling pathway |
| cg08234897 | RHEB | TSS200 | mTOR signaling pathway |
| cg08914916 | MRPS12 | TSS200 | Ribosome |
| cg09455096 | XPO1 | TSS200 | Nucleocytoplasmic transport, Ribosome biogenesis in eukaryotes |
| cg11149452 | GRB2 | TSS200 | mTOR signaling pathway |
| cg11343579 | RPS15 | TSS200 | Ribosome |
| cg11491989 | RAN | TSS200 | Nucleocytoplasmic transport, Ribosome biogenesis in eukaryotes |
| cg12052497 | LPIN2 | TSS200 | mTOR signaling pathway |
| cg13515774 | RPTOR | TSS200 | mTOR signaling pathway |
| cg13774825 | AKT1 | TSS1500 | mTOR signaling pathway |
| cg13954297 | CDK1 | TSS200 | Cell cycle |
| cg14073058 | MRPL30 | TSS200 | Ribosome |
| cg14360865 | RNF152 | TSS200 | mTOR signaling pathway |
| cg14493980 | RPL23 | TSS200 | Ribosome |
| cg15361065 | WNT5A | TSS1500 | mTOR signaling pathway |
| cg15931557 | PIK3R3 | TSS200 | mTOR signaling pathway |
| cg15988263 | IPO13 | TSS200 | Nucleocytoplasmic transport |
| cg16711450 | INS | TSS1500 | mTOR signaling pathway |
| cg16911228 | CDKN2C | TSS1500 | Cell cycle |
| cg17629447 | CHUK | TSS200 | mTOR signaling pathway |
| cg17678877 | MRPS9 | TSS200 | Ribosome |
| cg18152712 | E2F3 | TSS1500 | Cell cycle |
| cg19423197 | ATP6V1A | TSS1500 | mTOR signaling pathway |
| cg19987840 | SPATA5 | TSS200 | Ribosome biogenesis in eukaryotes |
| cg22682751 | CDC25A | TSS1500 | Cell cycle |
| cg23022344 | SRRM1 | TSS200 | Nucleocytoplasmic transport |
| cg23239078 | CDKN1C | TSS1500 | Cell cycle |
| cg23369529 | GADD45A | TSS1500 | Cell cycle |
| cg23823879 | SAP18 | TSS200 | Nucleocytoplasmic transport |
| cg24642820 | NUP210 | TSS1500 | Nucleocytoplasmic transport |
| cg24719901 | RSL24D1 | TSS200 | Ribosome |
| cg25092540 | NUP188 | TSS1500 | Nucleocytoplasmic transport |
| cg25245636 | ANAPC11 | TSS1500 | Cell cycle |
| cg25503001 | XPO4 | TSS200 | Nucleocytoplasmic transport |
| cg25988717 | POM121L2 | TSS1500 | Nucleocytoplasmic transport |
| cg26345046 | CCNA1 | TSS1500 | Cell cycle |
| cg26545834 | SENP2 | TSS200 | Nucleocytoplasmic transport |

**Supplemental Table 3. Pathway enrichment for genes with high negative DNAm and gene expression correlation.** Pathway databases, pathway names, and adjusted p-value are reported in the table. Pathways with FDR-adjusted p-values less than 0.05 are statistically significant.

| ID | Name | Source | p-value | q-value FDR B&H | Hit Count in Query List | Hit Count in Genome |
| --- | --- | --- | --- | --- | --- | --- |
| 83022 | Terpenoid backbone biosynthesis | BioSystems: KEGG | 1.06E-04 | 1.72E-02 | 8 | 22 |
| 83105 | Pathways in cancer | BioSystems: KEGG | 1.12E-04 | 1.72E-02 | 50 | 395 |
| 1404799 | Endocrine resistance | BioSystems: KEGG | 5.84E-04 | 4.56E-02 | 17 | 96 |
| 1269825 | Mitotic Anaphase | BioSystems: REACTOME | 2.54E-07 | 4.21E-04 | 34 | 179 |
| 1269823 | Mitotic Metaphase and Anaphase | BioSystems: REACTOME | 2.91E-07 | 4.21E-04 | 34 | 180 |
| 1269820 | Mitotic Prometaphase | BioSystems: REACTOME | 3.60E-07 | 4.21E-04 | 25 | 111 |
| 1269741 | Cell Cycle | BioSystems: REACTOME | 5.43E-07 | 4.77E-04 | 80 | 622 |
| 1269763 | Cell Cycle, Mitotic | BioSystems: REACTOME | 7.66E-07 | 5.38E-04 | 69 | 515 |
| 1269821 | Resolution of Sister Chromatid Cohesion | BioSystems: REACTOME | 1.23E-06 | 6.39E-04 | 23 | 103 |
| 1270038 | Regulation of cholesterol biosynthesis by SREBP (SREBF) | BioSystems: REACTOME | 1.27E-06 | 6.39E-04 | 16 | 55 |
| 1269826 | Separation of Sister Chromatids | BioSystems: REACTOME | 1.63E-06 | 7.14E-04 | 31 | 168 |
| 1269519 | RHO GTPases Activate Formins | BioSystems: REACTOME | 1.20E-05 | 3.25E-03 | 23 | 117 |
